# Assessing eligibility for lung cancer screening: Parsimonious multi-country ensemble machine learning models for lung cancer prediction

**DOI:** 10.1101/2023.01.27.23284974

**Authors:** Thomas Callender, Fergus Imrie, Bogdan Cebere, Nora Pashayan, Neal Navani, Mihaela van der Schaar, Sam M Janes

**Author notes:** Corresponding Author: Dr Thomas Callender, Department of Respiratory Medicine, University College London, 5 University Street, London, WC1E 6JF. Joint senior authors (Mihaela van der Schaar, Sam M Janes).

## Abstract

**Background:** Ensemble machine learning could support the development of highly parsimonious prediction models that maintain the performance of more complex models whilst maximising simplicity and generalisability, supporting the widespread adoption of personalised screening. In this work, we aimed to develop and validate ensemble machine learning models to determine eligibility for risk-based lung cancer screening.

**Methods:** For model development, we used data from 216,714 ever-smokers in the UK Biobank prospective cohort and 26,616 high-risk ever-smokers in the control arm of the US National Lung Screening randomised controlled trial. We externally validated our models amongst the 49,593 participants in the chest radiography arm and amongst all 80,659 ever-smoking participants in the US Prostate, Lung, Colorectal and Ovarian Screening Trial (PLCO). Models were developed to predict the risk of two outcomes within five years from baseline: diagnosis of lung cancer, and death from lung cancer. We assessed model discrimination (area under the receiver operating curve, AUC), calibration (calibration curves and expected/observed ratio), overall performance (Brier scores), and net benefit with decision curve analysis.

**Results:** Models predicting lung cancer death (UCL-D) and incidence (UCL-I) using three variables – age, smoking duration, and pack-years – achieved or exceeded parity in discrimination, overall performance, and net benefit with comparators currently in use, despite requiring only one-quarter of the predictors. In external validation in the PLCO trial, UCL-D had an AUC of 0.803 (95% CI: 0.783-0.824) and was well calibrated with an expected/observed (E/O) ratio of 1.05 (95% CI: 0.95-1.19). UCL-I had an AUC of 0.787 (95% CI: 0.771-0.802), an E/O ratio of 1.0 (0.92-1.07). The sensitivity of UCL-D was 85.5% and UCL-I was 83.9%, at 5-year risk thresholds of 0.68% and 1.17%, respectively 7.9% and 6.2% higher than the USPSTF-2021 criteria at the same specificity.

**Conclusions:** We present parsimonious ensemble machine learning models to predict the risk of lung cancer in ever-smokers, demonstrating a novel approach that could simplify the implementation of risk-based lung cancer screening in multiple settings.

## Introduction

Screening, early detection, and disease prevention programmes are increasingly bespoke, with risk prediction algorithms determining an individual’s eligibility and management.^1–3^ Such personalisation promises to improve the benefit-to-harm profile of such interventions and ultimately health outcomes.^4–6^ However, the delivery of these programmes at a population scale requires two conditions of risk prediction models: that they generalise well to contexts where there are insufficient data for model development, retraining, or validation; and, that the trade-off between model complexity and implementation feasibility is considered. In this work, we couple state-of-the-art ensemble machine learning and multi-country data to explicitly maximise model parsimony and generalisability, an approach that holds promise in multiple disease areas.

Screening for lung cancer – the foremost cause of death from cancer worldwide^7^ – with low-dose computed tomography (LDCT) has been associated with a 20-24% reduction in lung cancer-specific mortality amongst those at high risk.^8,9^ However, the ideal method to identify those at high risk remains unresolved. The US Preventive Services Taskforce (USPSTF) recommends the use of risk-factors – age, pack-years smoked, and quit-years for former smokers – to select screening participants.^10^ Nevertheless, identifying individuals for lung cancer screening based on risk prediction models has been shown to have both better benefit-to-harm profiles and cost-effectiveness than using risk factors alone,^11–14^ leading to risk-model-based selection criteria in European lung cancer screening pilots.^15^

To date, most externally validated prediction models for lung cancer have been developed in US datasets,^12,16–21^ reflecting the relatively limited availability of suitable cohorts with long-term follow-up for prognostic modelling. This implies that most global healthcare systems that implement risk-based lung cancer screening will use prediction models developed in a US population, often using variables such as ethnicity, whose categorisation varies between countries and individual datasets, and academic qualifications that differ both over time and between jurisdictions. In the UK, existing models have been shown to underperform in specific groups, such as the more socio-economically deprived, where underestimation of risk could lead to a screening programme systematically widening health inequalities.^22^

Furthermore, the risk models currently in use are a challenge to implement. In the UK, eligibility for lung cancer screening pilots is based on the PLCOm2012 and Liverpool Lung Project risk models, requiring 19 variables, few of which are routinely available.^23^ Collecting these variables from an individual who is potentially eligible and explaining the results currently averages between five and ten minutes. To determine the screening eligibility of one million people would therefore require up to 87 full-time staff a whole year, presenting a formidable obstacle to an effective national screening programme.

In this study, we hypothesized that using ensemble machine learning with training data spanning different geographic regions, populations, and average risk levels, we could develop predictive models for lung cancer screening with a minimum number of features that has broad applicability. In so doing, we aimed to combine the simplicity of risk-factor-based criteria with the improved predictive performance of risk models, whilst maintaining generalisability to new settings.

## Methods

### Data sources and study population

#### Development and internal validation datasets

For model development, we first used data on 216,714 ever-smokers without a prior history of lung cancer from the UK Biobank^24^ before creating a multi-country dataset that combined UK Biobank and US National Lung Screening Trial (NLST)^8^ data (n=26,616) (Figure 1 and eFigures 1-2 in the Supplement). We selected the NLST because it is geographically distinct, includes a higher risk cohort, and has greater ethnic diversity than the UK Biobank.

**Figure 1:**
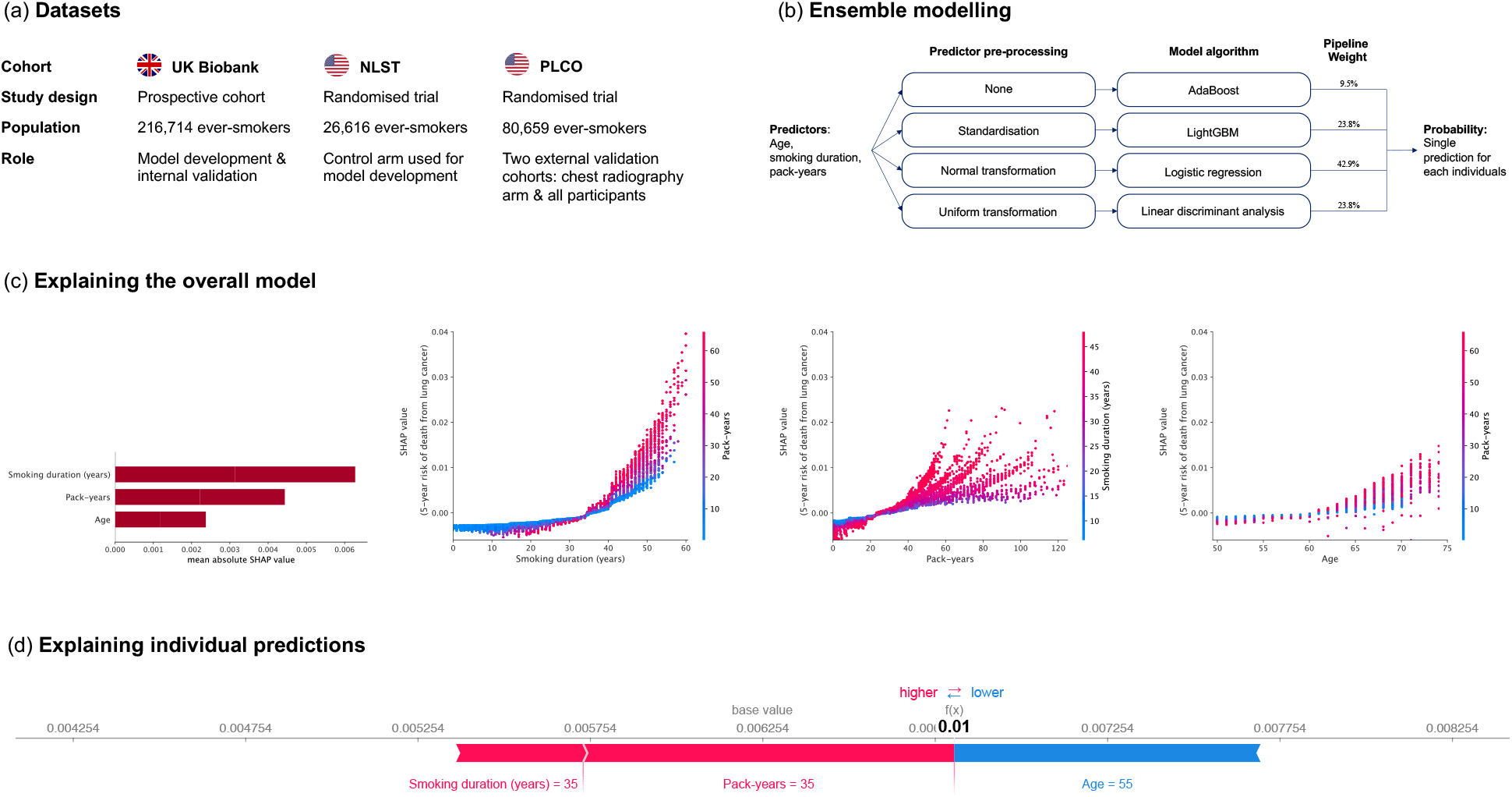
Developing the UCL models to determine lung cancer screening eligibility. A multi-country dataset comprising the UK Biobank and NLST was used to develop new models before external validation in the PLCO chest radiography arm (allowing benchmark comparison with existing models developed in the PLCO control arm) and the full PLCO cohort. The ensemble modelling approach involves optimising individual modelling pipelines before combining their results as a single prediction for each individual. (b) shows details of the UCL-D model, including the weights attributed to each pipeline in generating a single prediction for the five-year risk of lung cancer for any individual. (c) shows the contribution of different variables to overall predictions as well as interactions between predictors, analysed using Shapely Additive Explanations (SHAP).^32^ The first subfigure in (c) shows that smoking duration was the most important variable when making predictions of an individual’s risk of dying from lung cancer, followed by pack-years smoked, and finally age. The three subsequent dependence subplots show the relationship between the predictor (x-axis) against the outcome (y-axis) – the importance of knowing that predictor value when making a prediction. The vertical dispersion shows the degree of interaction effects present, whilst the colour corresponds to a second variable. The plots show that smoking for less than approximately 35 years had relatively little impact on model predictions, with a steep inflection and increasing interaction between smoking duration and pack-years after this point. Interestingly, in the subsequent subfigure showing the relationship between pack-years and lung cancer death, we see that there are distinct clusters of individuals based on their smoking duration projecting as a fan. This relationship between smoking duration and pack-years mirrors that seen in the previous sub-figure, with duration trumping quantity of cigarettes smoked unless both are high. In other words, those individuals who smoke for short periods of time have a lower predicted risk, even if they smoke relatively large quantities. This reflects our understanding of lung biology and the ability of the lung to repair itself if an individual stops smoking.^52^ Lastly amongst subfigures of (c) we see that age has relatively limited impact on the model under the age of 60. In (d), we explain an individual at the proposed risk threshold (0.68% five-year risk of death from lung cancer) for this model. Relative to the average, this individual’s predicted probability is lowered by their age (55) but raised by their smoking duration and pack-years, leading to a predicted probability above the average for this dataset. This can provide a useful check on the model and improve trustworthiness. Further details can be found in the Appendix.

#### External validation datasets

For model validation, we used data from 40,593 ever-smokers without a prior history of lung cancer from the chest radiography arm of the U.S. Prostate, Lung, Colorectal and Ovarian Cancer Screening (PLCO)^25^ trial (eFigure 3 in the Supplement). This allowed benchmarking against comparator models that were developed in the control arm of the PLCO trial. Chest radiography was found to have no impact on lung cancer mortality, nor a statistically significant impact on lung cancer incidence.^25^ In sensitivity analyses presented in the Appendix, we report model performance in the full PLCO dataset (n=80,659).

### Missing data

We used multiple imputation by chained equations (MICE) with predictive mean matching to generate imputed development and validation datasets.^26^ We generated 10 imputed sets of the UK Biobank and NLST, based on an average missingness amongst candidate predictors in the UK Biobank of 11%. As missingness was <1% for all relevant variables in the PLCO, we created five imputed PLCO datasets. See Appendix (Table S1, eFigures S4-6) for further details.

### Outcomes

We developed models to predict the absolute cumulative risk of two outcomes within five years from baseline: diagnosis of lung cancer, and death from lung cancer. Lung cancer status and primary cause of death in the UK Biobank were determined by linked national cancer registry and Office for National Statistics data.^24^ In the NLST and PLCO, lung cancer diagnosis and primary cause of death were confirmed by review of medical records and death certificates, respectively.^25,27^

### Model development

We developed ensembles of machine learning pipelines using AutoPrognosis, open-source automated machine learning software.^28,29^ In this analysis, AutoPrognosis was used to optimise pipelines consisting of a variable pre-processing step followed by model selection and training. These optimised pipelines were subsequently combined and a single prediction for any individual generated by a weighted combination of the predictions made by each of the four pipelines independently, with weighting by Bayesian model averaging (Figure 1).^30^ We trialled model algorithms including logistic regression, random forests and state-of-the-art Gradient Boosting approaches (see eMethods, eFigures 7-8, and eTables S2-3). Throughout, pipelines were trained and selected to maximise model discrimination, measured with the area under the receiver operating curve (AUC).

### Model explanation

We used the Kernel Shapely Additive Explanations (SHAP)^31^ algorithm for model explanation and analysis of predictor interactions (Figure 1). Kernel SHAP is a permutation-based method theoretically based in coalitional game theory. In summary, each variable is passed to a model one-by-one, with the change in predictions that occurs attributed to this model.^32,33^ Further details are available in the Appendix.

### Variable selection

For pragmatic reasons, we considered candidate predictors from the UK Biobank that were also present in the NLST and PLCO. We settled on our final list of predictors based on the literature, domain expertise, variable distributions, generalisability to multiple settings, and model discrimination in the UK Biobank.

### Statistical analysis

We considered a model’s overall performance with the Brier score,^34^ discrimination with the area under the receiver operating curve (AUC), calibration with calibration curves and the ratio of expected-to-observed cases, and clinical usefulness with decision curve analysis.^35^ Calibration curves were calculated by splitting individuals into ten risk deciles based on their predicted risk before compared predicted probability against observed risk, the latter calculated using a Kaplan-Meier model. For a measure of clinical utility, we considered the net benefit of models across a range of risk thresholds.^35^ We compared model discrimination with a two-tailed bootstrap test using the methods of Hanley and MacNeil, modified by Robin and colleagues.^36,37^ To determine potential risk thresholds for our models, we used a fixed population strategy, comparing the number of individuals eligible for screening in the entire PLCO external validation dataset using the USPSTF-2021 criteria.

In both internal and external validation, we generated 1,000 bootstrap resamples with replacement for all analyses; central estimates and 95% confidence intervals were calculated with the percentile method. We used optimism-corrected metrics for internal validation. All analyses were conducted with R^38^ and Python^39^.

### Model comparisons

For benchmark comparisons, we compared our new models to the USPSTF-2021 criteria (age 50-80, ≥20 pack-year smoking history, and quit within the last 15 years if a former smoker),^10^ as well as existing risk models that are either in use (PLCOm2012^18^ and Liverpool Lung Project (LLP) version 2^40^) or have been externally validated and consistently shown to outperform other risk models (the Lung Cancer Death Risk Assessment Tool [LCDRAT] and Lung Cancer Risk Assessment Tool [LCRAT]^19^) (eTable 4 in the Supplement).^13,22,41,42^ All comparator models predict the five-year risk of death (LCDRAT) or developing lung cancer (LCRAT, LLP) except for the PLCOm2012 which predicts the six-year risk of lung cancer occurrence. A third, recalibrated, version of LLP has been developed. Because it is not currently in use, we present full comparative analyses in the Appendix but note that in using the same predictors and coefficients as LLP version 2, its discrimination is equivalent. Further, we also compared against Cox models developed using the same dataset (see eMethods), and the constrained versions of the LCDRAT, LCRAT, and PLCOm2012 models.

All variables were available for comparator models except the LLP. For the LLP, in the UK Biobank, data were not available for age at which a family member developed lung cancer. Following ten Haaf and colleagues,^41^ and reflecting UK lung cancer epidemiology,^43^ we assumed that all with a family history of lung cancer were aged over 60. In the PLCO dataset, asbestos exposure and prior history of pneumonia were not available and were set to zero. We used the lcmodels package in R to calculate predictions for the PLCOm2012, LCRAT and LCDRAT models.^44^

### Code and model availability

To facilitate use of the UCL models, we have developed a website and have made the models themselves available (https://github.com/callta/lung_cancer_risk_models) as a package. The underlying code for AutoPrognosis is available from https://github.com/vanderschaarlab/AutoPrognosis.

## Results

The descriptive characteristics of the UK Biobank and NLST development datasets, and the PLCO external validation dataset are presented in Table 1. Characteristics by outcome are presented in eTables 5-8 in the Appendix. The number of cancers diagnosed and deaths from lung cancer are presented by follow-up period in eTable 9.

**Table 1:** Descriptive characteristics of development and validation cohorts.

| Characteristic | Development cohorts |  | Validation cohort |
| --- | --- | --- | --- |
|  | UK Biobank<br>n=216,714 | NLST controls<br>n=26,616 | PLCO radiography<br>arm n=49,593 |
| Age (n, %) |  |  |  |
| <50 | 43,170 (19.92) | - | - |
| 50-54 | 30,077 (13.88) | - | - |
| 55-59 | 39,539 (18.24) | 11,384 (42.77) | 13,965 (34.41) |
| 60-64 | 57,295 (26.44) | 8,170 (30.7) | 12,623 (31.1) |
| 65-69 | 45,520 (21.0) | 4,741 (17.81) | 9,117 (22.46) |
| ≥70 | 1,113 (0.51) | 2,321 (8.72) | 4,879 (12.02) |
| Missing | 0 (0.0) | 0 (0.0) | 9 (0.02) |
| Sex – Female (n, %) | 103,698 (47.85) | 10,919 (41.02) | 16,892 (41.61) |
| Missing | 0 (0.0) | 0 (0.0) | 0 (0.0) |
| Ethnicity – White (n, %) | 208,255 (96.47) | 24,165 (91.50) | 35,818 (88.29) |
| Missing | 830 (0.38) | 206 (0.77) | 23 (0.06) |
| Highest qualification (n, %) |  |  |  |
| Degree | 59,705 (28.07) | 8,213 (31.03) | 13,149 (32.44) |
| Some college | 16,501 (7.76) | 6,072 (22.94) | 9,434 (23.27) |
| Post-secondary school | 33,588 (15.79) | 10,100 (38.17) | 14,403 (35.53) |
| Secondary school | 57,646 (27.11) | 1,211 (4.58) | 3,083 (7.61) |
| None of the above | 45,231 (21.27) | 868 (3.28) | 464 (1.14) |
| Missing | 4043 (1.87) | 152 (0.57) | 60 (0.15) |
| Household income (GBP £) |  |  |  |
| <18,000 | 49,067 (26.45) | - | - |
| 18,000-30,999 | 49,023 (26.42) | - | - |
| 31,000-51,999 | 46,120 (24.86) | - | - |
| 52,000-100,000 | 33,020 (17.8) | - | - |
| >100,000 | 8,296 (4.47) | - | - |
| Missing | 31,188 (14.39) | - | - |
| Body mass index |  |  |  |
| <18.5 | 1,084 (0.50) | 240 (0.91) | 310 (0.77) |
| 18.5-24 | 62,715 (29.1) | 7,302 (27.65) | 12,743 (31.78) |
| 25-29 | 94,272 (43.75) | 11,442 (43.33) | 17,280 (43.1) |
| 30-34 | 41,469 (19.24) | 5,219 (19.76) | 7,035 (17.55) |
| ≥35 | 15,954 (7.40) | 2,205 (8.35) | 2,726 (6.80) |
| Missing | 1,220 (0.56) | 208 (0.78) | 499 (1.23) |
| Smoking status |  |  |  |
| Former | 164,714 (76.01) | 13,764 (51.71) | 8,073 (19.89) |
| Current | 52,000 (23.99) | 12,852 (48.29) | 32,520 (80.11) |
| Missing | 0 (0.0) | 0 (0.0) | 0 (0.0) |
| Pack-years of smoking (n, %) |  |  |  |
| <10 | 35,222 (23.59) | 0 (0.0) | 6,609 (16.63) |
| 11-19 | 39,914 (26.73) | 0 (0.0) | 7,605 (19.13) |
| 20-29 | 29,471 (19.74) | 4 (0.02) | 5,839 (14.69) |
| 30-39 | 20,596 (13.79) | 6,865 (25.79) | 5,108 (12.85) |
| ≥40 | 24,125 (16.16) | 19,747 (74.19) | 14,592 (36.71) |
| Missing | 67,386 (31.09) | 0 (0.0) | 840 (2.07) |
| Personal history of cancer (n, %) | 19,386 (8.95) | 1,197 (4.5) | 1,837 (4.53) |
| Missing | 0 (0.0) | 0 (0.0) | 5 (0.01) |
| COPD / Emphysema / Bronchitis (n, %) | 6,616 (3.06) | 4,617 (17.35) | 3,617 (8.91) |
| Missing | 454 (0.21) | 0 (0.0) | 0 (0.0) |
| Family history of lung cancer (n, %) | 28,765 (13.52) | 5,734 (21.54) | 4,566 (11.71) |
| Missing | 3,944 (1.82) | 0 (0.0) | 1602 (3.95) |

Abbreviations: GBP, British Pounds; COPD, Chronic Obstructive Pulmonary Disease.

We found that age, smoking duration (years), and pack-years of smoking, drove most predictions. This led us to focus our analyses on developing two models: UCL-D and UCL-I, that used just these three variables. UCL-D predicts the five-year risk of dying from lung cancer and was a weighted ensemble consisting of four modelling algorithms: AdaBoost^45,46^, LightGBM^47^, Logistic Regression and Linear Discriminant Analysis. UCL-I predicts the five-year risk of developing lung cancer and included AdaBoost^45,46^, LightGBM^47^, Bagging, and CatBoost^48^ algorithms. Details of the ensemble pipelines, their weightings and algorithm hyperparameters are presented in the Appendix (eFigures 7-8 and eTable S2-3). Using an ensemble approach led to higher discrimination than equivalent Cox models (eTable 10).

### UCL models

In internal and external validation, UCL-D and UCL-I showed good discrimination (Table 2), overall performance (Appendix Table S11), and calibration (Figure 2), both overall and across subgroups. In external validation in the PLCO radiography arm, UCL-D had an AUC of 0.803 (95% CI: 0.783-0.824), an expected/observed (E/O) ratio of 1.05 (95% CI: 0.95-1.19), and a Brier score of 0.0084 (95% CI: 0.0075-0.0093). UCL-I had an AUC of 0.787 (95% CI: 0.771-0.802), an E/O ratio of 1.0 (0.92-1.07), and a Brier score of 0.0153 (0.0142-0.0164).

**Table 2:** Discriminative accuracy (AUC) overall and by subgroup in the UK Biobank and PLCO radiography cohorts.

|  | Risk of death from lung cancer |  | Risk of developing lung cancer |  |  |  |
| --- | --- | --- | --- | --- | --- | --- |
|  | UCL-D | LCDRAT | UCL-I | LCRAT | PLCOm2012 | LLPv2 |
| **UK Biobank** |  |  |  |  |  |  |
| Overall | 0.826 (0.815-0.838) | 0.829 (0.813-0.841) | 0.810 (0.800-0.820) | 0.815 (0.805-0.827) | 0.797 (0.783-0.81) | 0.779 (0.767-0.793) |
| Age category |  |  |  |  |  |  |
| 40-49 | 0.747 (0.659-0.838) | 0.755 (0.616-0.904) | 0.781 (0.727-0.834) | 0.793 (0.692-0.865) | 0.797 (0.721-0.876) | 0.672 (0.575-0.775) |
| 50-59 | 0.807 (0.780-0.834) | 0.803 (0.769-0.834) | 0.777 (0.754-0.799) | 0.781 (0.751-0.808) | 0.779 (0.751-0.81) | 0.719 (0.687-0.748) |
| 60-72 | 0.788 (0.772-0.802) | 0.792 (0.769-0.805) | 0.769 (0.756-0.781) | 0.776 (0.762-0.791) | 0.765 (0.750-0.780) | 0.740 (0.725-0.754) |
| Sex |  |  |  |  |  |  |
| Female | 0.830 (0.812-0.846) | 0.825 (0.798-0.844) | 0.812 (0.798-0.825) | 0.811 (0.793-0.831) | 0.796 (0.780-0.817) | 0.771 (0.750-0.791) |
| Male | 0.820 (0.805-0.838) | 0.829 (0.808-0.845) | 0.809 (0.796-0.821) | 0.819 (0.802-0.831) | 0.798 (0.781-0.815) | 0.783 (0.767-0.797) |
| Smoking status |  |  |  |  |  |  |
| Former | 0.815 (0.796-0.833) | 0.813 (0.792-0.834) | 0.794 (0.780-0.808) | 0.798 (0.783-0.816) | 0.778 (0.760-0.798) | 0.775 (0.757-0.794) |
| Current | 0.773 (0.751-0.793) | 0.780 (0.759-0.802) | 0.778 (0.763-0.792) | 0.787 (0.773-0.801) | 0.767 (0.751-0.781) | 0.743 (0.726-0.757) |
| Ethnicity |  |  |  |  |  |  |
| Other | 0.818 (0.722-0.982) | 0.806 (0.631-0.972) | 0.810 (0.740-0.889) | 0.789 (0.660-0.862) | 0.827 (0.755-0.905) | 0.798 (0.737-0.857) |
| White | 0.825 (0.812-0.837) | 0.827 (0.813-0.840) | 0.809 (0.799-0.819) | 0.815 (0.805-0.827) | 0.796 (0.781-0.809) | 0.778 (0.765-0.791) |
| Household income |  |  |  |  |  |  |
| <18,000 | 0.786 (0.764-0.802) | 0.791 (0.768-0.811) | 0.769 (0.755-0.785) | 0.782 (0.762-0.800) | 0.766 (0.747-0.785) | 0.742 (0.722-0.759) |
| 18,000 to 30,999 | 0.816 (0.791-0.837) | 0.812 (0.787-0.836) | 0.794 (0.777-0.814) | 0.803 (0.781-0.822) | 0.785 (0.762-0.805) | 0.749 (0.722-0.771) |
| 31,000 to 51,999 | 0.811 (0.780-0.848) | 0.822 (0.772-0.861) | 0.791 (0.764-0.816) | 0.788 (0.752-0.824) | 0.771 (0.733-0.807) | 0.757 (0.719-0.799) |
| 52,000 to 100,000 | 0.836 (0.789-0.877) | 0.828 (0.763-0.883) | 0.821 (0.785-0.853) | 0.808 (0.755-0.852) | 0.798 (0.741-0.851) | 0.790 (0.736-0.835) |
| >100,000 | 0.744 (0.614-0.938) | 0.756 (0.536-0.924) | 0.808 (0.733-0.876) | 0.772 (0.634-0.875) | 0.738 (0.583-0.849) | 0.755 (0.624-0.850) |
| **PLCO radiography arm** |  |  |  |  |  |  |
| Overall | 0.803 (0.783-0.824) | 0.811 (0.793-0.829) | 0.787 (0.771-0.802) | 0.798 (0.784-0.814) | 0.792 (0.779-0.808) | 0.743 (0.726-0.762) |
|  | UCL-D | LCDRAT | UCL-I | LCRAT | PLCOm2012 | LLPv2 |
| Age category |  |  |  |  |  |  |
| 55-59 | 0.800 (0.745-0.844) | 0.815 (0.766-0.858) | 0.797 (0.762-0.833) | 0.817 (0.778-0.847) | 0.794 (0.756-0.825) | 0.729 (0.695-0.767) |
| 60-64 | 0.793 (0.753-0.831) | 0.799 (0.764-0.830) | 0.759 (0.722-0.790) | 0.776 (0.742-0.804) | 0.770 (0.741-0.796) | 0.716 (0.678-0.751) |
| 65-69 | 0.787 (0.747-0.823) | 0.806 (0.768-0.840) | 0.781 (0.752-0.809) | 0.792 (0.765-0.823) | 0.798 (0.775-0.823) | 0.747 (0.715-0.777) |
| 70-74 | 0.747 (0.694-0.790) | 0.725 (0.673-0.773) | 0.728 (0.685-0.768) | 0.723 (0.677-0.760) | 0.720 (0.682-0.753) | 0.675 (0.628-0.715) |
| Sex |  |  |  |  |  |  |
| Female | 0.800 (0.771-0.828) | 0.801 (0.771-0.831) | 0.771 (0.745-0.796) | 0.784 (0.760-0.804) | 0.784 (0.764-0.805) | 0.731 (0.699-0.757) |
| Male | 0.803 (0.773-0.828) | 0.818 (0.791-0.841) | 0.795 (0.774-0.814) | 0.807 (0.789-0.825) | 0.798 (0.776-0.819) | 0.755 (0.731-0.779) |
| Smoking status |  |  |  |  |  |  |
| Former | 0.813 (0.787-0.842) | 0.819 (0.793-0.843) | 0.791 (0.768-0.814) | 0.802 (0.781-0.824) | 0.793 (0.774-0.814) | 0.741 (0.715-0.768) |
| Current | 0.681 (0.642-0.721) | 0.705 (0.667-0.744) | 0.677 (0.650-0.717) | 0.698 (0.672-0.736) | 0.694 (0.669-0.724) | 0.651 (0.622-0.696) |
| Qualifications |  |  |  |  |  |  |
| Degree | 0.680 (0.323-0.921) | 0.709 (0.427-0.898) | 0.610 (0.455-0.779) | 0.681 (0.551-0.799) | 0.629 (0.509-0.751) | 0.609 (0.493-0.742) |
| Some college | 0.756 (0.654-0.834) | 0.796 (0.688-0.900) | 0.750 (0.686-0.818) | 0.771 (0.698-0.848) | 0.726 (0.640-0.803) | 0.663 (0.597-0.737) |
| Post-secondary | 0.730 (0.632-0.825) | 0.772 (0.651-0.859) | 0.753 (0.688-0.826) | 0.780 (0.718-0.843) | 0.763 (0.704-0.827) | 0.741 (0.670-0.814) |
| Secondary school | 0.638 (0.542-0.719) | 0.650 (0.55-0.742) | 0.620 (0.545-0.691) | 0.644 (0.578-0.710) | 0.664 (0.608-0.718) | 0.643 (0.582-0.707) |
| None of above | 0.700 (0.671-0.725) | 0.699 (0.675-0.728) | 0.673 (0.651-0.697) | 0.689 (0.667-0.710) | 0.693 (0.670-0.714) | 0.644 (0.621-0.667) |
| Ethnicity |  |  |  |  |  |  |
| Asian | 0.839 (0.805-0.874) | 0.857 (0.825-0.888) | 0.804 (0.770-0.834) | 0.823 (0.791-0.848) | 0.820 (0.787-0.848) | 0.752 (0.714-0.784) |
| Black | 0.805 (0.754-0.847) | 0.802 (0.751-0.842) | 0.804 (0.771-0.835) | 0.811 (0.778-0.841) | 0.800 (0.764-0.828) | 0.748 (0.709-0.790) |
| Other | 0.791 (0.754-0.822) | 0.789 (0.757-0.818) | 0.765 (0.737-0.793) | 0.774 (0.749-0.802) | 0.765 (0.741-0.791) | 0.728 (0.700-0.755) |
| White | 0.734 (0.671-0.795) | 0.744 (0.675-0.805) | 0.735 (0.696-0.779) | 0.741 (0.697-0.785) | 0.755 (0.715-0.791) | 0.707 (0.658-0.758) |

**Figure 2:**
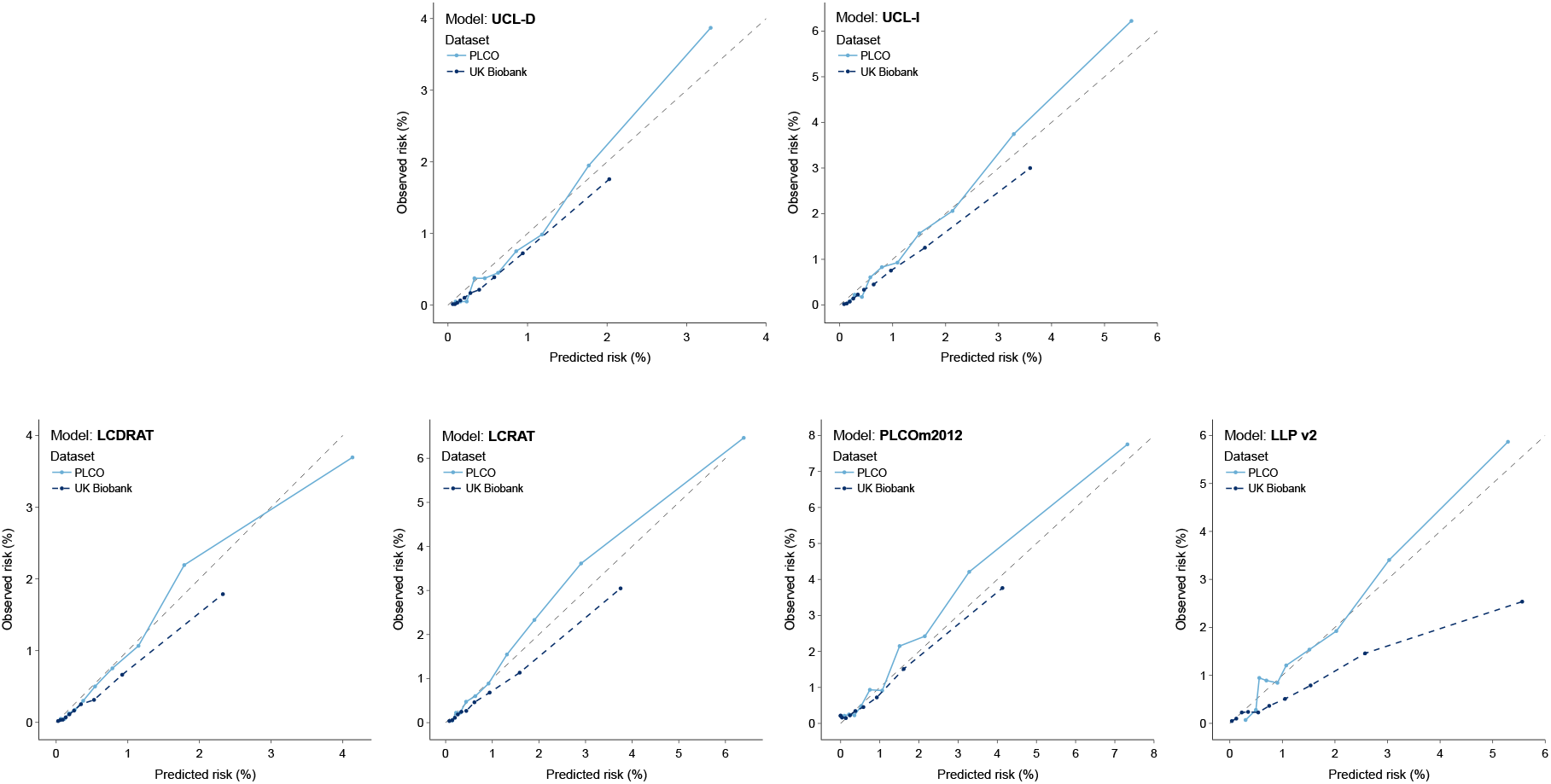
Calibration curves. Calibration curves showing UCL and comparator models in the UK Biobank (dark blue dashed lines) and US PLCO radiography arm (light blue line). Curves were generated by splitting individuals into ten risk deciles based on their predicted risk. Each curve shows the mean predicted risk against the observed risk by risk decile. Observed risk was calculated using a Kaplan-Meier estimator. UCL models showed good calibration in external validation in the PLCO intervention arm, particularly at predicted risk between 1-2% at which risk thresholds are commonly set. At these thresholds there was modest underprediction with the LCDRAT, LCRAT, and PLCOm2012 models in the PLCO intervention arm. All models modestly overpredicted risk in the UK Biobank, with the exception of the Liverpool Lung Project (LLP) version 2 model, which strongly overpredicted risk.

### Discrimination

Despite using approximately one-quarter of the variables, UCL-D achieved parity in discrimination with the LCDRAT (AUC: 0.811, 95%: 0.793-0.829, *p*=0.18 for difference with UCL-D). UCL-I achieved parity with PLCOm2012 (AUC: 0.792, 0.779-0.808, *p*=0.15 for difference in AUCs) and showed greater discrimination than LLP versions 2 and 3 (*p*<0.001).

### Calibration

The UCL models were well calibrated across risk thresholds at which eligibility for screening is typically set, tending modestly towards underprediction in the highest risk decile in the PLCO radiography arm (Figure 2). By contrast, PLCOm2012 and LCRAT tended modestly towards underprediction at deciles corresponding to observed risks of 1-4%, which is more clinically disadvantageous than overprediction. As the PLCOm2012, LCDRAT and LCRAT models were developed in the control arm of the PLCO trial, the strong relative performance of the UCL models is notable. All models modestly overpredicted risk in the UK Biobank cohort, with the extent of overprediction most notable for the LLP version 2.

### Overall performance

When considering Brier scores, an overall measure of model performance comparing the closeness of predicted probabilities and observed outcomes,^49^ there was little or no distinction between the models in the UK Biobank and PLCO radiography arm (Appendix Table S11). In the PLCO radiography arm, both models predicting the five-year risk of death, UCL-D and LCDRAT had a Brier score of 0.0084 (95% CI: 0.0075-0.0093). Brier scores vary with prevalence; consequently, models predicting the risk of developing lung cancer had higher scores. Nevertheless, the same pattern was observed: UCL-I had a Brier score of 0.0153 (95% CI: 0.0142-0.0164), LCRAT a score of 0.0152 (95% CI: 0.0143-0.0164), and LLP version 2 a score of 0.0153 (95% CI: 0.0143-0.0165).

### Risk thresholds to select individuals for screening

Using the USPSTF-2021 criteria, 34,654 (43.0%) of the entire PLCO dataset would be eligible for lung cancer screening. All UCL models had higher sensitivity than the USPSTF-2021 at an equivalent specificity, with the gains in sensitivity higher when predicting five-year risk of death from lung cancer (eTable 12). For UCL-I at a five-year risk threshold of 1.17%, the gains in sensitivity were 6.2% relative to the USPSTF-2021 criteria (83.9% [95% CI: 82.0-86.1%] vs 77.7% [95% CI: 75.8-80.2%]). By contrast, UCL-D at a five-year risk threshold of 0.68% would lead to a 7.9% increase in sensitivity (85.5% [95% CI: 82.8-88.2%] vs 77.5% [95% CI: 74.6-80.9%]) for the same specificity.

At the aforementioned risk cut-offs, 96.2% of individuals selected by UCL-D would also have been eligible for screening with UCL-I. By 10-years of follow-up, those selected for screening with UCL-D but not UCL-I tended towards a greater risk of developing and dying from lung cancer than those selected by UCL-I but not UCL-D, though this trend was not statistically significant (eFigure 9; Logrank test: *p*=0.15 for differences in lung cancer deaths and *p*=0.41 for differences in lung cancers).

### Clinical usefulness

Using decision curve analysis, at all risk thresholds, the net benefit of the UCL models is greater than screening using the USPSTF-2021 criteria (Figure 3 and eFigure 10). At suggested risk thresholds, the net benefit of compared risk models other than LLP are equivalent.

**Figure 3:**
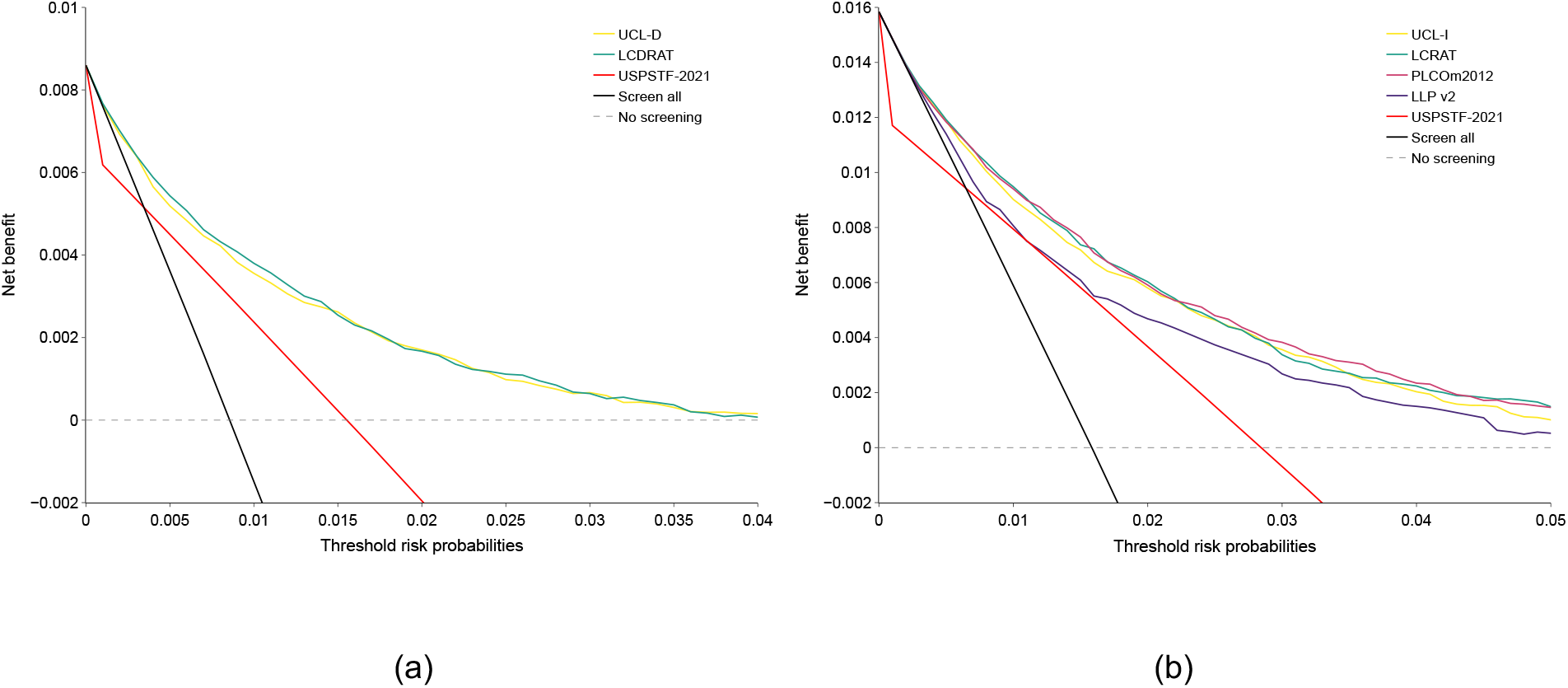
Decision curves of selected models in the PLCO validation cohort. Net benefit across a range of thresholds of models predicting 5-year risk of death from lung cancer (A) and developing lung cancer (B) compared against USPSTF-2021 screening eligibility criteria in the PLCO intervention arm validation dataset. The PLCOm2012 model predicts six-year risk of lung cancer. As the performance of PLCOm2012 over a five-year timeframe was similar to that of six-years, for comparability, predictions over a five-year timeframe are shown here. All models studied except the Liverpool Lung Project (LLP) version 2 had a greater net clinical benefit than using the USPSTF-2021 criteria for screening eligibility across all risk thresholds. All other risk models had a comparable net benefit to each other.

## Discussion

We have developed parsimonious models for lung cancer screening that combine the simplicity of existing risk factor-based criteria with the predictive performance of complex risk prediction models. Unique amongst existing risk prediction models for lung cancer screening, we have combined large United Kingdom and United States cohorts to train our models on over 240,000 individuals’ data with differing risk levels, to improve the generalisability of our models. Furthermore, we show in benchmarking comparisons that ensemble machine learning models with three predictors – age, smoking duration, and smoking pack-years – have equivalent predictive performance and clinical usefulness to existing models requiring eleven predictors.

In the UK, eligibility for National Health Service screening pilots is based on meeting either a five-year absolute risk of lung cancer of ≥2.5% with the LLP risk score or a six-year absolute risk of ≥1.51% with the PLCOm2012.^23^ The use of two risk scores where eligibility differs by more than a percentage point in predicted absolute risk, and where a higher risk is tolerated over a five-year period than a six-year period, highlights the policy challenge in adopting the optimal risk-based approach for a particular setting. This approach requires the collection of 19 different predictors, as well as the mapping of US educational levels and US ethnicity categorisations to the UK. With an estimated seven million current smokers in the UK^50^ – even ignoring former smokers – the time and resource requirements to determine screening eligibility at a population scale will be challenging. Using three unambiguous variables but with equivalent or improved performance, the UCL models could be completed more easily online or in primary healthcare, simplifying the implementation of lung cancer screening.

In keeping with Katki and colleagues,^19^ we found that UCL-D, predicting the risk of death from lung cancer, had greater discrimination than models predicting lung cancer occurrence. In these analyses, there was >96% overlap between UCL-D and UCL-I in terms of those selected for screening, with those selected by UCL-D but not UCL-I showing a trend towards a greater risk of death from lung cancer with longer follow-up (eFigure 9). In microsimulation modelling, overall outcomes differed little between a model predicting death from lung cancer compared with models predicting developing lung cancer.^13^ Given this, UCL-D would be the more appropriate model to consider for implementation.

In this analysis, we used ensemble machine learning to leverage the predictions of several optimised model pipelines. Ensemble modelling is based on the concept that different models make different types of mistake, and their errors begin to cancel each other out, such that combining these statistical models could be expected to improve the performance that any one might achieve.^51^ By iteratively trialling and optimising a wide range of state-of-the-art modelling approaches before subsequently creating ensembles of these approaches, AutoPrognosis ensures that the strongest performing model for that dataset will be derived and allows reproducibility by transparently showing how models were selected. This avoids the need to develop multiple independent models.

This study has several limitations. We have used retrospective data, such that findings may differ if used to prospectively determine screening eligibility. However, both the PLCOm2012 and the LLP models have been studied in prospective settings, establishing the benefits of risk-model against risk-factor-based screening. By benchmarking against these models, we can be confident in the performance of our models in a screening programme. To confirm the generalisability of our models, validation in datasets from beyond the US and UK will be the subject of further work. Finally, our risk models exclude never-smokers. To date, no risk model has been able to discriminate those never smokers with sufficient risk to meet existing criteria for lung cancer screening.

In summary, we have developed prognostic models to determine lung cancer screening eligibility that require only three variables – age, smoking duration, and pack-years – that perform at or above parity with existing risk models in use. Further validation in alternative datasets as well as prospective implementation should be considered.

## Data Availability

UK Biobank, NLST, and PLCO data were used on license (references 68073, NLST-806 and PLCO-801, respectively). These data cannot be shared directly, however, researchers can apply for these data from the UK Biobank and the US National Institutes of Health.

## Acknowledgments

This research has been conducted using the UK Biobank Resource under application number 68073 and we wish to thank all participants in the included studies, as well as the National Cancer Institute for access to NCI’s data collected by the National Lung Screening Trial (NLST) and Prostate, Lung, Colorectal and Ovarian (PLCO) Cancer Screening Trial. The statements contained herein are solely those of the authors and do not represent or imply concurrence or endorsement by NCI. We also wish to thank Arjun Nair and Sujin Kang for their feedback on earlier versions of this project, as well as Stephen Duffy for his comments on this work.

## Contributor Statement

TC conceived the study with SJ and MvdS. TC developed the models and performed the analyses. All authors provided critical input into model development. TC drafted the first draft of the manuscript; all authors were involved in manuscript revision. TC and SJ had full access to the data in the study and can take responsibility for the integrity of the data and the accuracy of the data analysis.

## Ethics

The University College London Research Ethics Committee gave ethical approval for this study (reference: 19131/001).

