## Supplementary Appendix for "Assessing eligibility for lung cancer screening: Parsimonious multi-country ensemble machine learning models for lung cancer prediction"

**Authors and affiliations:**

Thomas Callender[1], Fergus Imrie[2], Bogdan Cebere[3], Nora Pashayan[4], Neal Navani[1], Mihaela van der Schaar[3,5,6*], Sam M Janes[1*]

[1] Department of Respiratory Medicine, 5 University Street, University College London
[2] Department of Electrical and Computer Engineering, University of California, Los Angeles
[3] Department of Applied Mathematics and Theoretical Physics, University of Cambridge
[4] Department of Applied Health Research, 1-19 Torrington Place, University College London
[5] Cambridge Centre for AI in Medicine, University of Cambridge
[6] Alan Turing Institute

*Joint senior authors

### Participant flow diagrams

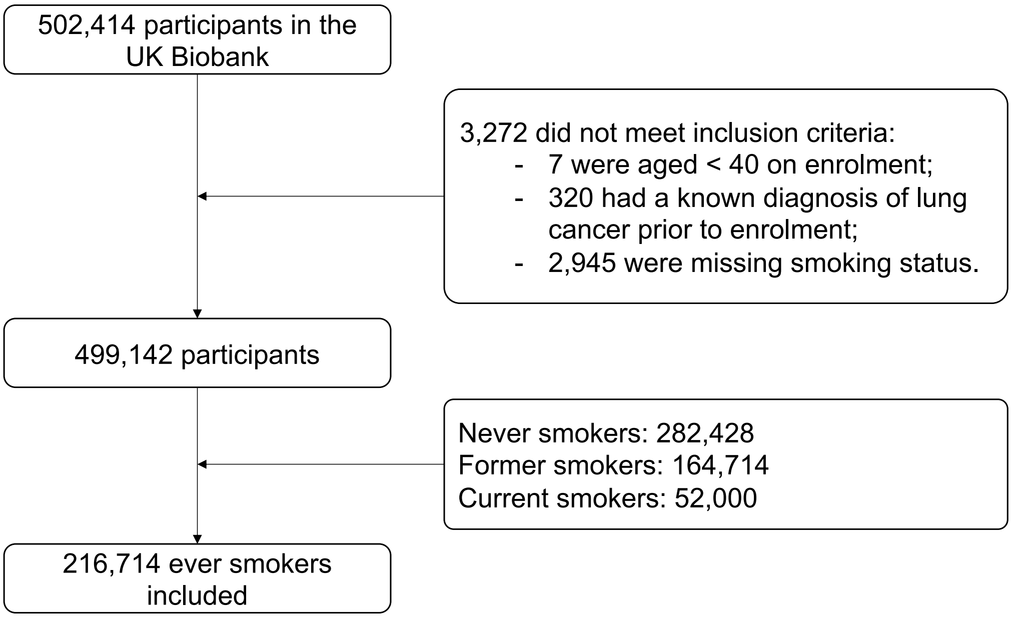

**eFigure 1:** Flow diagram of UK Biobank participants

From the UK Biobank, we included 216,714 ever-smoking individuals without a known diagnosis of lung cancer (ICD-10 codes C33-C34) aged 40 or more at baseline (eFigure 1). The UK Biobank is a prospective cohort of men and women recruited between 2006-2010 from 22 assessment centres across the UK which combines phenotypical data with ongoing linkage to national cancer and mortality registries.[1] During this timeframe, the UK has not had a systematic screening programme for lung cancer.

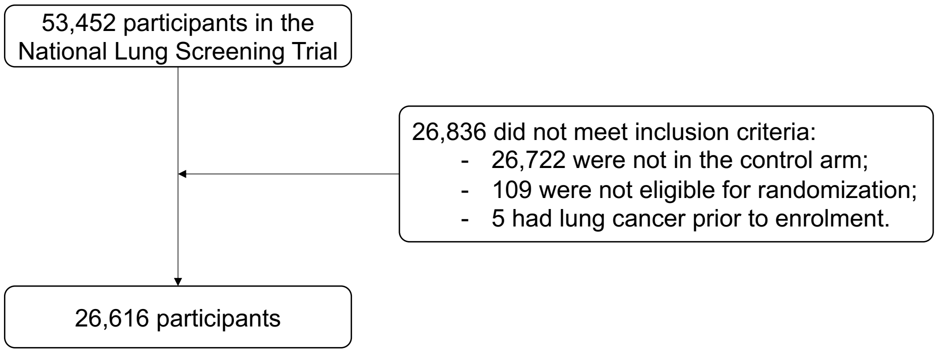

**eFigure 2:** Flow diagram of NLST participants included

From the NLST, we included the 26,616 individuals without a prior history of lung cancer who had been randomised to the control arm of the trial (eFigure 2). The NLST was a randomised controlled trial of lung cancer screening comparing computed tomography (CT) against chest radiography in 33 US centres between 2002-2004 with follow-up through 2009.[2] Participation in the NLST was restricted to those considered at high risk of developing lung cancer: a 30 pack-year smoking history and, if a former smoker, to have quit within 15 years of enrolment.[2]

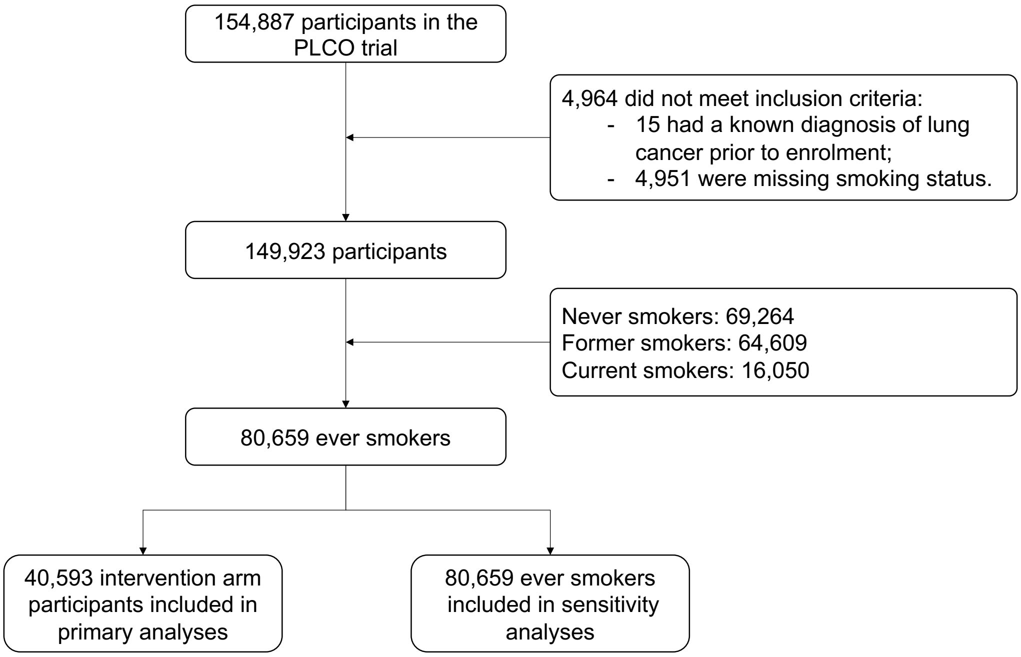

**eFigure 3:** Flow diagram of PLCO participants included

The PLCO was a randomised controlled trial of lung cancer screening with chest radiography compared to a non-interventional control that took place between 1993-2001 in 10 US centres.[3]

### Variable recoding, missing data, and multiple imputation

#### UK Biobank: Recoding smoking variables

To determine smoking status in the UK Biobank, we used their self-recorded smoking status (field 20116). We then re-coded the 9,010 participants who had declared (field 1249) they had never tried smoking, had smoked only once or twice, or less than 100 cigarettes in their lifetime (field 2644) to non-smokers. The final numbers by smoking status were: 282,428 never smokers, 165,714 former smokers and 52,000 current smokers.

#### UK Biobank: Missing data & multiple imputation

As the most influential risk factor for lung cancer, we specifically analysed patterns of missingness in smoking variables. Three smoking variables were key: age start smoking, age stop smoking, and smoking intensity (number of cigarettes smoked per day). Based on these three variables, variables such as smoking duration and pack-years are calculated.

In the UK Biobank development cohort, just over two-thirds of all participants (68.9%), and over five-sixths of those who developed lung cancer (86.3%) and of those who died from lung cancer (86.8%) during follow-up had complete data across all included predictors (eTable 1). However, we noted that missing all three smoking variables was the most common pattern amongst all participants, occurring in nearly one-quarter of ever-smokers in our development dataset (22.6%).

**eTable 1:** Distribution of complete data across smoking variables in our development cohort (UK Biobank)

| Variables missing | All participants, n=216,714 (n, %) | Developed lung cancer, n=3,449 (n, %) | Died from lung cancer, n=2,137 (n, %) |
|---|---|---|---|
| Complete data | 149,328 (68.91) | 2,977 (86.31) | 1,854 (86.76) |
| 1 | 10,264 (4.74) | 228 (6.61) | 137 (6.41) |
| 2 | 8,074 (3.73) | 56 (1.62) | 35 (1.64) |
| 3 | 49,048 (22.63) | 188 (5.45) | 111 (5.19) |

Within the context of prediction modelling, our interest is the relationship between missingness in a variable and the outcome – the informativeness of missingness – and the impact this has on predictive performance[4]. Amongst those with an outcome of interest, the most common pattern was to be missing one variable; missing one variable was not associated with a higher cumulative risk of developing lung cancer (log-rank test $p = 0.07$) or dying from lung cancer (log-rank test $p = 0.31$). By contrast, as shown in eFigure 5, missing three variables - the next most common pattern seen amongst those with an outcome of interest - was associated with a different risk of an outcome relative to those with complete data (log-rank test $p < 0.001$).

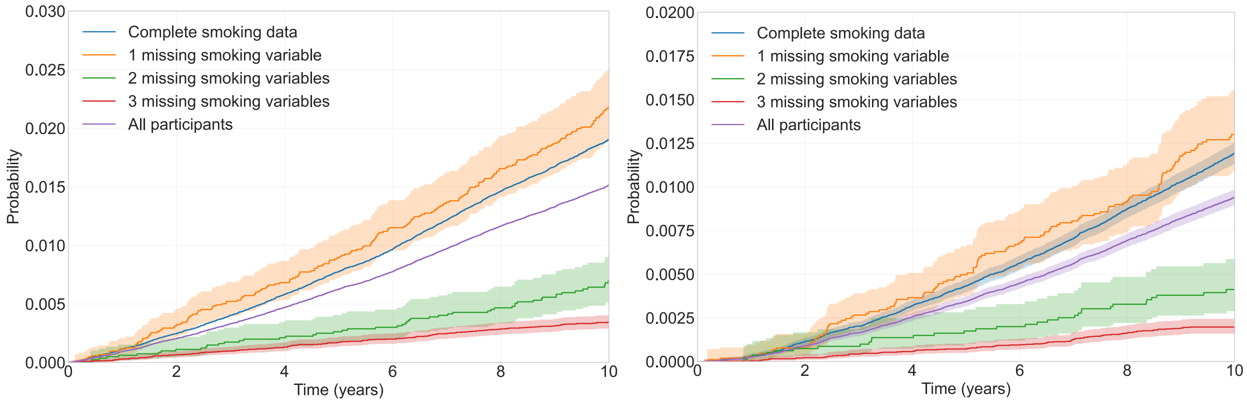

**eFigure 4:** Nelson-Aalen curves of probability of (a) developing and (b) dying from lung cancer amongst ever-smokers with different patterns of missing smoking data in the UK Biobank

Reviewing those participants who were missing all three key smoking variables, nearly all (99.9%, n=49,030) were recorded as former smokers who had only smoked occasionally (UK Biobank fields 20116 & 1249). These individuals were not questioned about the number of cigarettes they smoked per day. On removing these participants from analysis, the relationship between missingness and the outcomes of interest inverted (*p* < 0.001 for risk of developing or dying from lung cancer, eFigure 6).

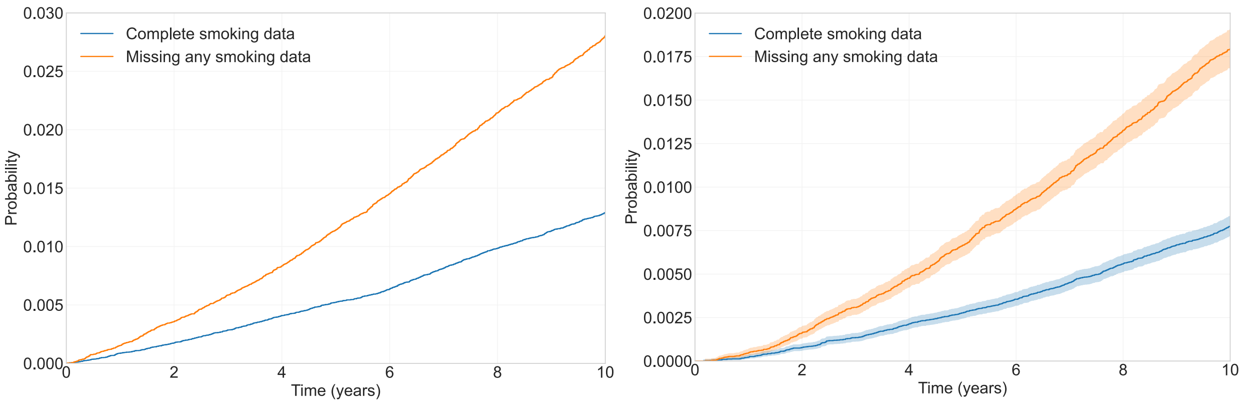

**eFigure 5:** Nelson-Aalen curves of probability of developing (a) and dying from (b) lung cancer amongst non-occasional ever-smokers not missing smoking intensity and duration in the UK Biobank

However, the former occasional smokers themselves showed different outcome profiles to both current smokers recorded to only smoke occasionally and non-smokers (eFigure 7). We therefore included them in our analyses.

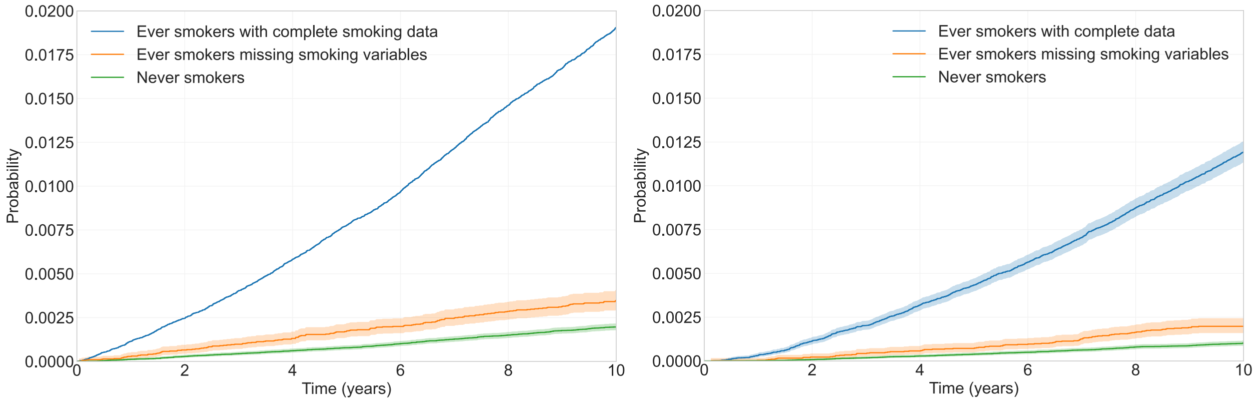

**eFigure 6:** Nelson-Aalen curves of probability of developing (a) and dying from (b) lung cancer amongst ever-smokers with smoking intensity and duration, ever-smokers missing smoking intensity and duration, and never-smokers in the UK Biobank

### UK Biobank Imputation

We generated 10 imputed datasets using multiple imputation by chained equations (MICE) and a tree-based Gradient Boosting imputation model (LightGBM) to avoid assumptions about statistical relationships between the variables, implemented with the Python package miceforest.[5] For each candidate predictor with missing data, a model was fit that consisted of both a common pool of candidate variables and lung cancer outcomes as well as a bespoke set of predictors that were specifically correlated with missingness in the variable of interest.[6]

To account for the systematic difference between former 'occasional' smokers missing cigarettes smoked per day and others, we imputed this number by taking a random draw from a *Gamma*(1.5626, 6.4) distribution. This equates to a median of 8 cigarettes smoked per *month* (interquartile range: 4-14, 2.5th and 97.5th centile: 0.8 and 31). We repeated this for each of the 10 imputed datasets.

Models were developed using a single imputed dataset as there are no established methods for pooling machine learning model hyperparameters between imputed datasets. However, to assess model performance, we pooled all ten imputed datasets using Rubin's rules.

### NLST & PLCO: Missing data and multiple imputation

Overall missingness was <1% for all relevant variables in both the NLST and PLCO datasets. For congruence when creating a multi-country dataset for model development, we created 10 imputed NLST datasets. Given the low level of missingness, we generated five imputed PLCO datasets. In both cases we used multiple imputation by chained equations with predictive mean matching, implemented as described above.

### Model development

We fit models using AutoPrognosis,[7,8] and for comparison and model validation with Cox proportional hazards regression.

In this analysis, AutoPrognosis searched for optimal pipelines – where each pipeline consists of three stages: dimensionality reduction, predictor pre-processing, and the model algorithm – from 252 potential combinations. The following algorithms were considered:

- Dimensionality reduction: none, variance thresholding, principal component analysis, independent component analysis.
- Predictor pre-processing: none, normalisation, polynomial interactions between features, scaling each predictor using its maximum absolute value, min-max scaling, uniform transformation, standardisation.
- Modelling algorithms: logistic regression, linear discriminant analysis, quadratic discriminant analysis, bagging, random forests, Adaboost[9–11], CatBoost[12,13], LightGBM[14,15], XGBoost[16,17].

AutoPrognosis uses Bayesian optimisation for pipeline selection,[7] whilst the hyperparameters of each modelling algorithm trialled were tuned using Optuna.[18] Ensembles are generated using both stacking and aggregating methods from the Python package combo[19] and by Bayesian model averaging.[7] All pipelines and ensembles were trained using five-fold cross-validation to maximise the area under the receiver operating curve (AUC), with the highest performing ensemble selected. We considered ensembles that consisted of up to four different modelling pipelines. In other words, should a single pipeline, for example one involving no dimensionality reduction, predictor standardisation, and subsequently the machine learning algorithm LightGBM, have had greater discrimination than an ensemble of several pipelines, this would have been selected.

#### Details of UCL-D

UCL-D is an ensemble of four modelling pipelines predicting the five-year risk of dying from lung cancer. The final ensemble, constituent pipelines, and the weighting assigned to each pipeline are shown in eFigure 7. The tuned hyperparameters for the AdaBoost and LightGBM algorithms are shown in eTable 2.

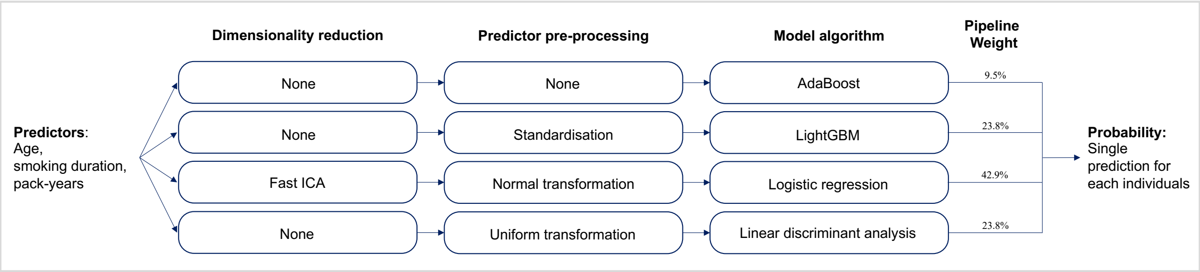

**eFigure 7:** The UCL-D ensemble and constituent pipelines (predicted outcome is five-year risk of death from lung cancer).

**eTable 2:** Hyperparameters for the AdaBoost and LightGBM machine learning algorithms in UCL-D

| Algorithm | Hyperparameter | Value |
|---|---|---|
| AdaBoost[9–11] | n_estimators | 10 |
| | learning_rate | 0.1 |
| LightGBM[14,15] | boosting_type | 'gbdt' |
| | learning_rate | 0.1 |
| | max_depth | 6 |
| | reg_alpha | 9.65x10-6 |
| | reg_lambda | 1.07x10-8 |
| | colsample_by_tree | 0.48 |
| | subsample | 0.61 |
| | num_leaves | 3 |
| | min_child_samples | 139 |

### Details of UCL-I

UCL-I is also an ensemble of four modelling pipelines that instead predicts the five-year risk of lung cancer occurrence. Details of the ensemble and tuned hyperparameters for the constituent machine learning algorithms are shown in eFigure 8 and eTable 3.

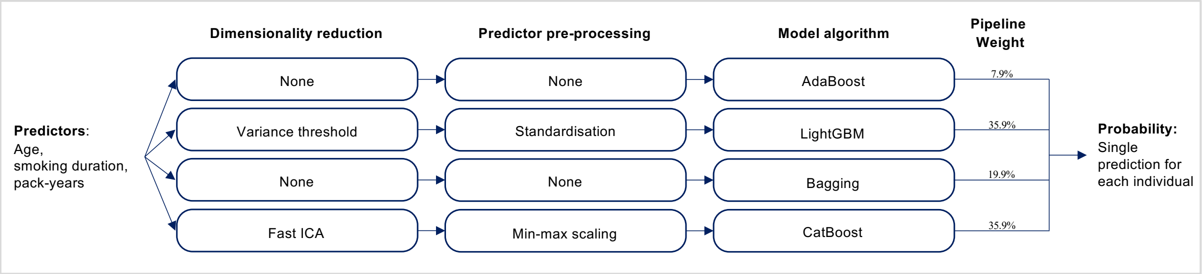

**eFigure 8:** Details of the UCL-I ensemble (predicted outcome is five-year risk of developing lung cancer).

**eTable 3:** Hyperparameters for the AdaBoost, LightGBM, and CatBoost machine learning algorithms in UCL-I

| Algorithm | Hyperparameter | Value |
|---|---|---|
| AdaBoost[9–11] | n_estimators | 10 |
| | learning_rate | 0.1 |
| LightGBM[14,15] | boosting_type | ‘gbdt’ |
| | learning_rate | 0.1 |
| | max_depth | 6 |
| | reg_alpha | 0.036 |
| | reg_lambda | 9.08x10-5 |
| | colsample_by_tree | 0.11 |
| | subsample | 0.11 |
| | num_leaves | 3 |
| | min_child_samples | 23 |
| CatBoost[12,13] | learning_rate | 0.039 |
| | depth | 7 |
| | l2_leaf_reg | 554.36 |
| | random_strength | 1.303 |
| | grow_policy | “Lossguide” |

### Cox models

We developed Cox models using the survival[20] and rms[21] packages in R. To account for non-linear relationships between age, smoking duration, pack-years and lung cancer (death), we used restricted cubic splines modelled with three knots. In common with both Tammemagi,[22] Katki,[23] and their colleagues, we found that no interactions between the three variables were significant within a Cox framework. Further, Cox models without including interaction terms had lower Akaike Information Criteria (AIC) values. We present the c-index for models both with and without interactions in the Appendix results.

**eTable 4:** Predictors used in comparator prognostic models

| Model | Predictors |
|---|---|
| UCL-D & UCL-I | 1. Age<br>2. Smoking duration (years)<br>3. Pack-years[a] |
| PLCOm2012[22] | 1. Age<br>2. Smoking status<br>3. Smoking duration (years)<br>4. Smoking intensity (number of cigarettes per day)<br>5. Quit-years (if former smoker)<br>6. Ethnicity (White, Black, Hispanic, Asian, American Indian or Alaskan Native, Native Hawaiian or Pacific Islander)<br>7. Highest qualification<br>8. Body-mass index<br>9. Chronic obstructive pulmonary disease (COPD)<br>10. Personal history of cancer<br>11. Family history of lung cancer |
| LCRAT & LCDRAT[23] | 1. Age<br>2. Sex<br>3. Ethnicity (White non-hispanic, Black non-Hispanic, Hispanic, Asian or other)<br>4. Highest qualification<br>5. Body-mass index<br>6. Smoking duration (years)<br>7. Smoking intensity (number of cigarettes per day)<br>8. Quit-years (if former smoker)<br>9. Pack-years<br>10. History of emphysema<br>11. Family history of lung cancer |
| LLP versions 2 & 3[24] | 1. Age<br>2. Sex<br>3. Smoking duration (years)<br>4. Pneumonia,<br>5. Asthma,<br>6. Bronchitis, emphysema, or COPD<br>7. Tuberculosis<br>8. Asbestos exposure<br>9. Personal history of cancer<br>10. Family history of lung cancer (none, before age 60, after age 70) |

[a]Pack-years combine smoking duration with the number of cigarettes smoked, with one pack-year equivalent to smoking 20 cigarettes per day for a year.

### Variable importance and interactions

We used Kernel Shapely Additive Explanations (SHAP) to disentangle the contribution of different variables to predictions, including interactions between variables.[25] Kernel SHAP is a feature attribution method designed to explain model predictions through the calculation of Shapely values, whose values represent the contribution made by individual predictors to the overall prediction.[25]

Shapely values are calculated by iteratively passing each predictor through a model and attributing changes in the predicted outcomes for an individual in its absence to the predictor. As an example, for an individual with an age of 55, smoking duration of 30 years and pack-years of 60, the change in prediction made if the model only used smoking duration and pack-years can be attributed to age. These attributions are subsequently averaged across the dataset to get the final Shapely values for each predictor.[26] With SHAP, the result is a linear model $\phi_0 + \sum \phi_i x_i$ ,[25] where $\phi_0$ is an intercept corresponding to the average predicted risk in the dataset being explained, along with the SHAP values $\phi_i$ for each of the predictors $x_i$ (i.e., age, smoking duration, and pack-years).

### Supplementary Results

**eTable 5: Descriptive characteristics of UK Biobank ever-smokers by outcome**

| | Lung cancer n=3,449 | No lung cancer n=213,265 | Lung cancer death n=2,137 | No lung cancer death n=214,577 | All participants n=216,714 |
|---|---|---|---|---|---|
| Age (n, %) | | | | | |
| <50 | 123 (3.57) | 43,047 (20.18) | 75 (3.51) | 43,095 (20.08) | 43,170 (19.92) |
| 50-54 | 243 (7.05) | 29,834 (13.99) | 152 (7.11) | 29,925 (13.95) | 30,077 (13.88) |
| 55-59 | 556 (16.12) | 38,983 (18.28) | 347 (16.24) | 39,192 (18.26) | 39,539 (18.24) |
| 60-64 | 1,180 (34.21) | 56,115 (26.31) | 712 (33.32) | 56,583 (26.37) | 57,295 (26.44) |
| 65-69 | 1,316 (38.16) | 44,204 (20.73) | 834 (39.03) | 44,686 (20.83) | 45,520 (21.0) |
| ≥70 | 31 (0.9) | 1,082 (0.51) | 17 (0.8) | 1,096 (0.51) | 1,113 (0.51) |
| *Missing* | 0 (0.0) | 0 (0.0) | 0 (0.0) | 0 (0.0) | 0 (0.0) |
| Sex - Female (n, %) | 1,574 (45.64) | 102,124 (47.89) | 936 (43.8) | 102,762 (47.89) | 103,698 (47.85) |
| *Missing* | 0 (0.0) | 0 (0.0) | 0 (0.0) | 0 (0.0) | 0 (0.0) |
| Ethnicity - White (n, %) | 3,368 (98.02) | 204,887 (96.44) | 2,097 (98.36) | 206,158 (96.45) | 208,255 (96.47) |
| *Missing* | 13 (0.38) | 817 (0.38) | 5 (0.23) | 825 (0.38) | 830 (0.38) |
| Highest qualification (n, %) | | | | | |
| Degree | 478 (14.3) | 59,227 (28.29) | 276 (13.29) | 59,429 (28.22) | 59,705 (28.07) |
| Some college | 310 (9.28) | 16,191 (7.73) | 200 (9.63) | 16,301 (7.74) | 16,501 (7.76) |
| Post-secondary school | 450 (13.46) | 33,138 (15.83) | 275 (13.24) | 33,313 (15.82) | 33,588 (15.79) |
| Secondary school | 725 (21.69) | 56,921 (27.19) | 437 (21.04) | 57,209 (27.17) | 57,646 (27.11) |
| None of the above | 1,379 (41.26) | 43,852 (20.95) | 889 (42.8) | 44,342 (21.06) | 45,231 (21.27) |
| *Missing* | 107 (3.1) | 3936 (1.85) | 60 (2.81) | 3983 (1.86) | 4043 (1.87) |
| In paid employment/self-employed (n, %) | 1,071 (31.16) | 115,762 (54.47) | 629 (29.49) | 116,204 (54.35) | 116,833 (54.1) |
| *Missing* | 12 (0.35) | 755 (0.35) | 4 (0.19) | 763 (0.36) | 767 (0.35) |
| Body mass index | | | | | |
| <18.5 | 46 (1.35) | 1,038 (0.49) | 29 (1.37) | 1,055 (0.49) | 1,084 (0.5) |
| 18.5-24 | 1,046 (30.65) | 61,669 (29.08) | 652 (30.78) | 62,063 (29.09) | 62,715 (29.1) |
| 25-29 | 1,437 (42.1) | 92,835 (43.77) | 904 (42.68) | 93,368 (43.76) | 94,272 (43.75) |
| 30-34 | 665 (19.48) | 40,804 (19.24) | 403 (19.03) | 41,066 (19.25) | 41,469 (19.24) |
| ≥35 | 219 (6.42) | 15,735 (7.42) | 130 (6.14) | 15,824 (7.42) | 15,954 (7.4) |
| *Missing* | 36 (1.04) | 1,184 (0.56) | 19 (0.89) | 1,201 (0.56) | 1,220 (0.56) |
| Household income (GBP £) | | | | | |
| <18,000 | 1,318 (47.6) | 47,749 (26.13) | 864 (50.2) | 48,203 (26.23) | 49,067 (26.45) |
| 18,000-30,999 | 790 (28.53) | 48,233 (26.39) | 492 (28.59) | 48,531 (26.4) | 49,023 (26.42) |
| 31,000-51,999 | 404 (14.59) | 45,716 (25.01) | 230 (13.36) | 45,890 (24.97) | 46,120 (24.86) |
| 52,000-100,000 | 208 (7.51) | 32,812 (17.95) | 110 (6.39) | 32,910 (17.9) | 33,020 (17.8) |
| >100,000 | 49 (1.77) | 8,247 (4.51) | 25 (1.45) | 8,271 (4.5) | 8,296 (4.47) |
| *Missing* | 680 (19.72) | 30,508 (14.31) | 416 (19.47) | 30,772 (14.34) | 31,188 (14.39) |
| | | | | | *Continued...* |
| Smoking status | | | | | |
| Former | 1,817 (52.68) | 162,897 (76.38) | 1,073 (50.21) | 163,641 (76.26) | 164,714 (76.01) |
| Current | 1,632 (47.32) | 50,368 (23.62) | 1,064 (49.79) | 50,936 (23.74) | 52,000 (23.99) |

### eTable 5: Descriptive characteristics of UK Biobank ever-smokers by outcome

| | Lung cancer n=3,449 | No lung cancer n=213,265 | Lung cancer death n=2,137 | No lung cancer death n=214,577 | All participants n=216,714 |
|---|---|---|---|---|---|
| *Missing* | 0 (0.0) | 0 (0.0) | 0 (0.0) | 0 (0.0) | 0 (0.0) |
| Age started smoking | | | | | |
| <16 | 1,223 (38.34) | 47,210 (30.34) | 792 (39.94) | 47,641 (30.38) | 48,433 (30.5) |
| 16-20 | 1,658 (51.97) | 87,974 (56.54) | 997 (50.28) | 88,635 (56.53) | 89,632 (56.45) |
| >20 | 309 (9.69) | 20,403 (13.11) | 194 (9.78) | 20,518 (13.09) | 20,712 (13.04) |
| *Missing* | 259 (7.51) | 57,678 (27.05) | 154 (7.21) | 57,783 (26.93) | 57,937 (26.73) |
| Years smoked | | | | | |
| <10 | 54 (1.7) | 16,910 (10.9) | 31 (1.57) | 16,933 (10.83) | 16,964 (10.71) |
| 10-19 | 176 (5.53) | 36,641 (23.62) | 105 (5.31) | 36,712 (23.48) | 36,817 (23.25) |
| 20-29 | 386 (12.14) | 38,039 (24.52) | 201 (10.16) | 38,224 (24.45) | 38,425 (24.27) |
| 30-39 | 787 (24.75) | 35,610 (22.95) | 480 (24.27) | 35,917 (22.97) | 36,397 (22.99) |
| ≥40 | 1,777 (55.88) | 27,954 (18.02) | 1,161 (58.7) | 28,570 (18.27) | 29,731 (18.78) |
| *Missing* | 269 (7.8) | 58,111 (27.25) | 159 (7.44) | 58,221 (27.13) | 58,380 (26.94) |
| Cigarettes per day (median, IQR) | | | | | |
| 1-10 | 547 (18.26) | 41,797 (28.4) | 338 (18.14) | 42,006 (28.32) | 42,344 (28.2) |
| 11-20 | 1,575 (52.57) | 76,880 (52.24) | 977 (52.44) | 77,478 (52.24) | 78,455 (52.24) |
| 21-30 | 545 (18.19) | 18,653 (12.67) | 336 (18.04) | 18,862 (12.72) | 19,198 (12.78) |
| 31-40 | 240 (8.01) | 6,837 (4.65) | 153 (8.21) | 6,924 (4.67) | 7,077 (4.71) |
| >40 | 89 (2.97) | 3,013 (2.05) | 59 (3.17) | 3,043 (2.05) | 3,102 (2.07) |
| *Missing* | 453 (13.13) | 66,085 (30.99) | 274 (12.82) | 66,264 (30.88) | 66,538 (30.7) |
| Pack-years of smoking (n, %) | | | | | |
| <10 | 155 (5.21) | 35,067 (23.96) | 91 (4.91) | 35,131 (23.82) | 35,222 (23.59) |
| 10-19 | 371 (12.46) | 39,543 (27.02) | 218 (11.76) | 39,696 (26.92) | 39,914 (26.73) |
| 20-29 | 538 (18.07) | 28,933 (19.77) | 321 (17.31) | 29,150 (19.77) | 29,471 (19.74) |
| 30-39 | 595 (19.99) | 20,001 (13.67) | 362 (19.53) | 20,234 (13.72) | 20,596 (13.79) |
| ≥40 | 1,318 (44.27) | 22,807 (15.58) | 862 (46.49) | 23,263 (15.77) | 24,125 (16.16) |
| *Missing* | 472 (13.69) | 66,914 (31.38) | 283 (13.24) | 67,103 (31.27) | 67,386 (31.09) |
| Personal history of cancer (n, %) | 515 (14.93) | 18,871 (8.85) | 291 (13.62) | 19,095 (8.9) | 19,386 (8.95) |
| *Missing* | 0 (0.0) | 0 (0.0) | 0 (0.0) | 0 (0.0) | 0 (0.0) |
| COPD / Emphysema / Bronchitis (n, %) | 409 (11.9) | 6,207 (2.92) | 248 (11.64) | 6,368 (2.97) | 6,616 (3.06) |
| *Missing* | 13 (0.38) | 441 (0.21) | 7 (0.33) | 447 (0.21) | 454 (0.21) |
| Family history of lung cancer (n, %) | 797 (23.72) | 27,968 (13.36) | 473 (22.78) | 28,292 (13.43) | 28,765 (13.52) |
| *Missing* | 89 (2.58) | 3,855 (1.81) | 61 (2.85) | 3,883 (1.81) | 3,944 (1.82) |

### eTable 6: Descriptive characteristics of NLST control ever-smokers by outcome

| | Lung cancer n=960 | No lung cancer n=25,656 | Lung cancer death n=545 | No lung cancer death n=26,071 | All participants n=26,616 |
|---|---|---|---|---|---|
| Age (n, %) | | | | | |
| 55-59 | 247 (25.73) | 11,137 (43.41) | 139 (25.5) | 11,245 (43.13) | 11,384 (42.77) |
| 60-64 | 299 (31.15) | 7,871 (30.68) | 165 (30.28) | 8,005 (30.7) | 8,170 (30.7) |
| 65-69 | 250 (26.04) | 4,491 (17.5) | 141 (25.87) | 4,600 (17.64) | 4,741 (17.81) |
| 70-74 | 164 (17.08) | 2,157 (8.41) | 100 (18.35) | 2,221 (8.52) | 2,321 (8.72) |
| *Missing* | 0 (0.0) | 0 (0.0) | 0 (0.0) | 0 (0.0) | 0 (0.0) |
| Sex – Female (n, %) | 390 (40.62) | 10,529 (41.04) | 212 (38.9) | 10,707 (41.07) | 10,919 (41.02) |
| *Missing* | 0 (0.0) | 0 (0.0) | 0 (0.0) | 0 (0.0) | 0 (0.0) |
| Ethnicity – White (n, %) | 871 (91.2) | 23,294 (91.51) | 492 (91.28) | 23,673 (91.5) | 24,165 (91.5) |
| *Missing* | 5 (0.52) | 201 (0.78) | 6 (1.1) | 200 (0.77) | 206 (0.77) |
| Qualifications (n, %) | | | | | |
| Degree | 214 (22.43) | 7,999 (31.36) | 116 (21.56) | 8,097 (31.23) | 8,213 (31.03) |
| Some college | 209 (21.91) | 5,863 (22.98) | 121 (22.49) | 5,951 (22.95) | 6,072 (22.94) |
| Post-secondary school | 430 (45.07) | 9,670 (37.91) | 247 (45.91) | 9,853 (38.0) | 10,100 (38.17) |
| Secondary school | 67 (7.02) | 1,144 (4.48) | 40 (7.43) | 1,171 (4.52) | 1,211 (4.58) |
| None of the above | 34 (3.56) | 834 (3.27) | 14 (2.6) | 854 (3.29) | 868 (3.28) |
| *Missing* | 6 (0.62) | 146 (0.57) | 7 (1.28) | 145 (0.56) | 152 (0.57) |
| Body mass index (n, %) | | | | | |
| <18.5 | 13 (1.37) | 227 (0.89) | 10 (1.86) | 230 (0.89) | 240 (0.91) |
| 18.5-24 | 339 (35.61) | 6,963 (27.35) | 193 (35.87) | 7,109 (27.48) | 7,302 (27.65) |
| 25-29 | 400 (42.02) | 11,042 (43.38) | 221 (41.08) | 11,221 (43.37) | 11,442 (43.33) |
| 30-34 | 144 (15.13) | 5,075 (19.94) | 88 (16.36) | 5,131 (19.83) | 5,219 (19.76) |
| ≥35 | 56 (5.88) | 2,149 (8.44) | 26 (4.83) | 2,179 (8.42) | 2,205 (8.35) |
| *Missing* | 8 (0.83) | 200 (0.78) | 7 (1.28) | 201 (0.77) | 208 (0.78) |
| Smoking status (n, %) | | | | | |
| Former | 362 (37.71) | 13,402 (52.24) | 189 (34.68) | 13,575 (52.07) | 13,764 (51.71) |
| Current | 598 (62.29) | 12,254 (47.76) | 356 (65.32) | 12,496 (47.93) | 12,852 (48.29) |
| *Missing* | 0 (0.0) | 0 (0.0) | 0 (0.0) | 0 (0.0) | 0 (0.0) |
| Age started smoking (n, %) | | | | | |
| <16 | 408 (42.5) | 9,524 (37.12) | 229 (42.02) | 9,703 (37.22) | 9,932 (37.32) |
| 16-20 | 471 (49.06) | 13,304 (51.86) | 268 (49.17) | 13,507 (51.81) | 13,775 (51.75) |
| >20 | 81 (8.44) | 2,828 (11.02) | 48 (8.81) | 2,861 (10.97) | 2,909 (10.93) |
| *Missing* | 0 (0.0) | 0 (0.0) | 0 (0.0) | 0 (0.0) | 0 (0.0) |
| Years smoked (n, %) | | | | | |
| <10 | 0 (0.0) | 0 (0.0) | 0 (0.0) | 0 (0.0) | 0 (0.0) |
| 10-19 | 1 (0.1) | 66 (0.26) | 0 (0.0) | 67 (0.26) | 67 (0.25) |
| 20-29 | 14 (1.46) | 1,749 (6.82) | 10 (1.83) | 1,753 (6.72) | 1,763 (6.62) |
| 30-39 | 196 (20.42) | 10,296 (40.13) | 110 (20.18) | 10,382 (39.82) | 10,492 (39.42) |
| ≥40 | 749 (78.02) | 13,545 (52.79) | 425 (77.98) | 13,869 (53.2) | 14,294 (53.7) |
| *Missing* | 0 (0.0) | 0 (0.0) | 0 (0.0) | 0 (0.0) | 0 (0.0) |
| | | | | | *Continued…* |

### eTable 6: Descriptive characteristics of NLST control ever-smokers by outcome

| | Lung cancer n=960 | No lung cancer n=25,656 | Lung cancer death n=545 | No lung cancer death n=26,071 | All participants n=26,616 |
|---|---|---|---|---|---|
| Cigarettes per day (n, %) | | | | | |
| <10 | 0 (0.0) | 0 (0.0) | 0 (0.0) | 0 (0.0) | 0 (0.0) |
| 10-19 | 42 (4.38) | 1,335 (5.2) | 17 (3.12) | 1,360 (5.22) | 1,377 (5.17) |
| 20-29 | 436 (45.42) | 12,321 (48.02) | 243 (44.59) | 12,514 (48.0) | 12,757 (47.93) |
| 30-39 | 221 (23.02) | 5,995 (23.37) | 129 (23.67) | 6,087 (23.35) | 6,216 (23.35) |
| ≥40 | 261 (27.19) | 6,005 (23.41) | 156 (28.62) | 6,110 (23.44) | 6,266 (23.54) |
| *Missing* | 0 (0.0) | 0 (0.0) | 0 (0.0) | 0 (0.0) | 0 (0.0) |
| Pack-years of smoking (n, %) | | | | | |
| <10 | 0 (0.0) | 0 (0.0) | 0 (0.0) | 0 (0.0) | 0 (0.0) |
| 10-19 | 0 (0.0) | 0 (0.0) | 0 (0.0) | 0 (0.0) | 0 (0.0) |
| 20-29 | 0 (0.0) | 4 (0.02) | 0 (0.0) | 4 (0.02) | 4 (0.02) |
| 30-39 | 112 (11.67) | 6,753 (26.32) | 59 (10.83) | 6,806 (26.11) | 6,865 (25.79) |
| ≥40 | 848 (88.33) | 18,899 (73.66) | 486 (89.17) | 19,261 (73.88) | 19,747 (74.19) |
| *Missing* | 0 (0.0) | 0 (0.0) | 0 (0.0) | 0 (0.0) | 0 (0.0) |
| Personal history of cancer (n, %) | 59 (6.15) | 1,138 (4.44) | 29 (5.32) | 1,168 (4.48) | 1,197 (4.5) |
| *Missing* | 0 (0.0) | 0 (0.0) | 0 (0.0) | 0 (0.0) | 0 (0.0) |
| COPD/Emphysema/Chronic bronchitis (n, %) | 267 (27.81) | 4,350 (16.96) | 131 (24.04) | 4,486 (17.21) | 4,617 (17.35) |
| *Missing* | 0 (0.0) | 0 (0.0) | 0 (0.0) | 0 (0.0) | 0 (0.0) |
| Family history of lung cancer (n, %) | 256 (26.67) | 5,478 (21.35) | 142 (26.06) | 5,592 (21.45) | 5,734 (21.54) |
| *Missing* | 0 (0.0) | 0 (0.0) | 0 (0.0) | 0 (0.0) | 0 (0.0) |

### eTable 7: Descriptive characteristics of PLCO radiography arm ever-smokers by outcome

| | Lung cancer n=1,734 | No lung cancer n=38,859 | Lung cancer death n=1,782 | No lung cancer death n=38,811 | All participants n=40,593 |
|---|---|---|---|---|---|
| Age (n, %) | | | | | |
| 55-59 | 357 (20.6) | 13,608 (35.03) | 382 (21.44) | 13,583 (35.01) | 13,965 (34.41) |
| 60-64 | 514 (29.66) | 12,109 (31.17) | 541 (30.36) | 12,082 (31.14) | 12,623 (31.1) |
| 65-69 | 548 (31.62) | 8,569 (22.06) | 560 (31.43) | 8,557 (22.05) | 9,117 (22.46) |
| ≥70 | 314 (18.12) | 4,565 (11.75) | 299 (16.78) | 4,580 (11.8) | 4,879 (12.02) |
| *Missing* | 1 (0.06) | 8 (0.02) | 0 (0.0) | 9 (0.02) | 9 (0.02) |
| Sex - Female (n, %) | 641 (36.97) | 16,251 (41.82) | 630 (35.35) | 16,262 (41.9) | 16,892 (41.61) |
| *Missing* | 0 (0.0) | 0 (0.0) | 0 (0.0) | 0 (0.0) | 0 (0.0) |
| Ethnicity - White (n, %) | 1,520 (87.66) | 34,298 (88.31) | 1,535 (86.19) | 34,283 (88.38) | 35,818 (88.29) |
| *Missing* | 0 (0.0) | 23 (0.06) | 1 (0.06) | 22 (0.06) | 23 (0.06) |
| Highest qualification (n, %) | | | | | |
| Degree | 402 (23.2) | 12,747 (32.85) | 424 (23.79) | 12,725 (32.84) | 13,149 (32.44) |
| Some college | 400 (23.08) | 9,034 (23.28) | 423 (23.74) | 9,011 (23.25) | 9,434 (23.27) |
| Post-secondary school | 679 (39.18) | 13,724 (35.37) | 685 (38.44) | 13,718 (35.4) | 14,403 (35.53) |
| Secondary school | 225 (12.98) | 2,858 (7.37) | 216 (12.12) | 2,867 (7.4) | 3,083 (7.61) |
| None of the above | 27 (1.56) | 437 (1.13) | 34 (1.91) | 430 (1.11) | 464 (1.14) |
| *Missing* | 1 (0.06) | 59 (0.15) | 0 (0.0) | 60 (0.15) | 60 (0.15) |
| Body mass index | | | | | |
| <18.5 | 18 (1.05) | 292 (0.76) | 22 (1.25) | 288 (0.75) | 310 (0.77) |
| 18.5-24 | 660 (38.57) | 12,083 (31.48) | 656 (37.19) | 12,087 (31.53) | 12,743 (31.78) |
| 25-29 | 708 (41.38) | 16,572 (43.18) | 739 (41.89) | 16,541 (43.15) | 17,280 (43.1) |
| 30-34 | 263 (15.37) | 6,772 (17.64) | 269 (15.25) | 6,766 (17.65) | 7,035 (17.55) |
| ≥35 | 62 (3.62) | 2,664 (6.94) | 78 (4.42) | 2,648 (6.91) | 2,726 (6.8) |
| *Missing* | 23 (1.33) | 476 (1.22) | 18 (1.01) | 481 (1.24) | 499 (1.23) |
| Smoking status | | | | | |
| Current | 772 (44.52) | 7,301 (18.79) | 818 (45.9) | 7,255 (18.69) | 8,073 (19.89) |
| Previous | 962 (55.48) | 31,558 (81.21) | 964 (54.1) | 31,556 (81.31) | 32,520 (80.11) |
| *Missing* | 0 (0.0) | 0 (0.0) | 0 (0.0) | 0 (0.0) | 0 (0.0) |
| Age started smoking | | | | | |
| <16 | 468 (27.29) | 7,357 (19.04) | 450 (25.57) | 7,375 (19.11) | 7,825 (19.39) |
| 16-20 | 978 (57.03) | 23,374 (60.49) | 1,029 (58.47) | 23,323 (60.43) | 24,352 (60.35) |
| >20 | 269 (15.69) | 7,907 (20.46) | 281 (15.97) | 7,895 (20.46) | 8,176 (20.26) |
| *Missing* | 19 (1.1) | 221 (0.57) | 22 (1.23) | 218 (0.56) | 240 (0.59) |
| Years smoked | | | | | |
| <10 | 23 (1.35) | 4,688 (12.3) | 17 (0.97) | 4,694 (12.33) | 4,711 (11.83) |
| 1-19 | 98 (5.75) | 7,788 (20.43) | 98 (5.61) | 7,788 (20.46) | 7,886 (19.81) |
| 20-29 | 184 (10.8) | 8,064 (21.16) | 206 (11.78) | 8,042 (21.12) | 8,248 (20.71) |
| 30-39 | 416 (24.41) | 9,068 (23.79) | 418 (23.91) | 9,066 (23.81) | 9,484 (23.82) |
| ≥40 | 983 (57.69) | 8,505 (22.32) | 1,009 (57.72) | 8,479 (22.27) | 9,488 (23.83) |
| *Missing* | 30 (1.73) | 746 (1.92) | 34 (1.91) | 742 (1.91) | 776 (1.91) |
| Cigarettes per day (n, %) | | | | | |

### eTable 7: Descriptive characteristics of PLCO radiography arm ever-smokers by outcome

| | Lung cancer n=1,734 | No lung cancer n=38,859 | Lung cancer death n=1,782 | No lung cancer death n=38,811 | All participants n=40,593 |
|---|---|---|---|---|---|
| 1-10 | 197 (11.41) | 10,237 (26.39) | 205 (11.54) | 10,229 (26.41) | 10,434 (25.76) |
| 11-20 | 611 (35.38) | 14,331 (36.95) | 636 (35.79) | 14,306 (36.93) | 14,942 (36.88) |
| 21-30 | 446 (25.83) | 7,503 (19.35) | 479 (26.96) | 7,470 (19.29) | 7,949 (19.62) |
| 31-40 | 297 (17.2) | 4,097 (10.56) | 279 (15.7) | 4,115 (10.62) | 4,394 (10.85) |
| >40 | 176 (10.19) | 2,616 (6.75) | 178 (10.02) | 2,614 (6.75) | 2,792 (6.89) |
| *Missing* | 7 (0.4) | 75 (0.19) | 5 (0.28) | 77 (0.2) | 82 (0.2) |
| Pack-years of smoking (n, %) | | | | | |
| <10 | 39 (2.3) | 6,570 (17.26) | 34 (1.95) | 6,575 (17.3) | 6,609 (16.63) |
| 10-19 | 114 (6.71) | 7,491 (19.69) | 120 (6.88) | 7,485 (19.69) | 7,605 (19.13) |
| 20-29 | 170 (10.01) | 5,669 (14.9) | 182 (10.44) | 5,657 (14.88) | 5,839 (14.69) |
| 30-39 | 168 (9.89) | 4,940 (12.98) | 164 (9.4) | 4,944 (13.01) | 5,108 (12.85) |
| ≥40 | 1,208 (71.1) | 13,384 (35.17) | 1,244 (71.33) | 13,348 (35.12) | 14,592 (36.71) |
| *Missing* | 35 (2.02) | 805 (2.07) | 38 (2.13) | 802 (2.07) | 840 (2.07) |
| Personal history of cancer (n, %) | 97 (5.59) | 1,740 (4.48) | 96 (5.39) | 1,741 (4.49) | 1,837 (4.53) |
| *Missing* | 0 (0.0) | 5 (0.01) | 0 (0.0) | 5 (0.01) | 5 (0.01) |
| COPD / Emphysema / Bronchitis (n, %) | 308 (17.76) | 3,309 (8.52) | 295 (16.55) | 3,322 (8.56) | 3,617 (8.91) |
| *Missing* | 0 (0.0) | 0 (0.0) | 0 (0.0) | 0 (0.0) | 0 (0.0) |
| Family history of lung cancer (n, %) | 292 (17.95) | 4,274 (11.44) | 297 (17.74) | 4,269 (11.44) | 4,566 (11.71) |
| *Missing* | 107 (6.17) | 1495 (3.85) | 108 (6.06) | 1494 (3.85) | 1602 (3.95) |

### eTable 8: Descriptive characteristics of all PLCO ever-smokers by outcome

| | Lung cancer n=3,356 | No lung cancer n=77,303 | Lung cancer death n=3,534 | No lung cancer death n=77,125 | All participants n=80,659 |
|---|---|---|---|---|---|
| Age (n, %) | | | | | |
| 55-59 | 692 (20.63) | 26,886 (34.79) | 743 (21.02) | 26,835 (34.8) | 27,578 (34.2) |
| 60-64 | 1,013 (30.19) | 24,095 (31.18) | 1,097 (31.04) | 24,011 (31.14) | 25,108 (31.14) |
| 65-69 | 1,035 (30.85) | 17,069 (22.09) | 1,107 (31.32) | 16,997 (22.04) | 18,104 (22.45) |
| ≥70 | 615 (18.33) | 9,233 (11.95) | 587 (16.61) | 9,261 (12.01) | 9,848 (12.21) |
| *Missing* | 1 (0.03) | 20 (0.03) | 0 (0.0) | 21 (0.03) | 21 (0.03) |
| Sex - Female (n, %) | 1,263 (37.63) | 32,484 (42.02) | 1,281 (36.25) | 32,466 (42.1) | 33,747 (41.84) |
| *Missing* | 0 (0.0) | 0 (0.0) | 0 (0.0) | 0 (0.0) | 0 (0.0) |
| Ethnicity - White (n, %) | 2,978 (88.74) | 68,252 (88.34) | 3,085 (87.32) | 68,145 (88.4) | 71,230 (88.36) |
| *Missing* | 0 (0.0) | 43 (0.06) | 1 (0.03) | 42 (0.05) | 43 (0.05) |
| Highest qualification (n, %) | | | | | |
| Degree | 777 (23.19) | 25,126 (32.59) | 838 (23.75) | 25,065 (32.58) | 25,903 (32.2) |
| Some college | 776 (23.16) | 17,978 (23.32) | 815 (23.09) | 17,939 (23.32) | 18,754 (23.31) |
| Post-secondary school | 1,341 (40.02) | 27,394 (35.53) | 1,393 (39.47) | 27,342 (35.54) | 28,735 (35.72) |
| Secondary school | 414 (12.35) | 5,726 (7.43) | 423 (11.99) | 5,717 (7.43) | 6,140 (7.63) |
| None of the above | 43 (1.28) | 877 (1.14) | 60 (1.7) | 860 (1.12) | 920 (1.14) |
| *Missing* | 5 (0.15) | 202 (0.26) | 5 (0.14) | 202 (0.26) | 207 (0.26) |
| Body mass index | | | | | |
| <18.5 | 44 (1.33) | 562 (0.74) | 50 (1.43) | 556 (0.73) | 606 (0.76) |
| 18.5-24 | 1,258 (38.03) | 24,084 (31.65) | 1,286 (36.9) | 24,056 (31.68) | 25,342 (31.91) |
| 25-29 | 1,387 (41.93) | 33,013 (43.38) | 1,471 (42.21) | 32,929 (43.37) | 34,400 (43.32) |
| 30-34 | 482 (14.57) | 13,249 (17.41) | 510 (14.63) | 13,221 (17.41) | 13,731 (17.29) |
| ≥35 | 137 (4.14) | 5,194 (6.83) | 168 (4.82) | 5,163 (6.8) | 5,331 (6.71) |
| *Missing* | 48 (1.43) | 1201 (1.55) | 49 (1.39) | 1200 (1.56) | 1249 (1.55) |
| Smoking status | | | | | |
| Former | 1,840 (54.83) | 62,769 (81.2) | 1,910 (54.05) | 62,699 (81.3) | 64,609 (80.1) |
| Current | 1,516 (45.17) | 14,534 (18.8) | 1,624 (45.95) | 14,426 (18.7) | 16,050 (19.9) |
| *Missing* | 0 (0.0) | 0 (0.0) | 0 (0.0) | 0 (0.0) | 0 (0.0) |
| Age started smoking | | | | | |
| <16 | 865 (26.07) | 14,583 (18.98) | 880 (25.21) | 14,568 (19.0) | 15,448 (19.27) |
| 16-20 | 1,936 (58.35) | 46,507 (60.52) | 2,058 (58.95) | 46,385 (60.5) | 48,443 (60.43) |
| >20 | 517 (15.58) | 15,755 (20.5) | 553 (15.84) | 15,719 (20.5) | 16,272 (20.3) |
| *Missing* | 38 (1.13) | 458 (0.59) | 43 (1.22) | 453 (0.59) | 496 (0.61) |
| Years smoked | | | | | |
| <10 | 51 (1.55) | 9,171 (12.12) | 48 (1.38) | 9,174 (12.15) | 9,222 (11.67) |
| 10-19 | 179 (5.43) | 15,358 (20.29) | 196 (5.65) | 15,341 (20.31) | 15,537 (19.67) |
| 20-29 | 344 (10.44) | 16,048 (21.2) | 386 (11.13) | 16,006 (21.19) | 16,392 (20.75) |
| 30-39 | 803 (24.37) | 18,008 (23.79) | 842 (24.28) | 17,969 (23.79) | 18,811 (23.81) |
| ≥40 | 1,918 (58.21) | 17,110 (22.6) | 1,996 (57.55) | 17,032 (22.55) | 19,028 (24.09) |
| *Missing* | 61 (1.82) | 1608 (2.08) | 66 (1.87) | 1603 (2.08) | 1669 (2.07) |
| | | | | | *Continued…* |

### eTable 8: Descriptive characteristics of all PLCO ever-smokers by outcome

| | Lung cancer n=3,356 | No lung cancer n=77,303 | Lung cancer death n=3,534 | No lung cancer death n=77,125 | All participants n=80,659 |
|---|---|---|---|---|---|
| Cigarettes per day (n, %) | | | | | |
| 1-10 | 379 (11.33) | 20,249 (26.26) | 404 (11.46) | 20,224 (26.29) | 20,628 (25.64) |
| 11-20 | 1,155 (34.54) | 28,180 (36.54) | 1,247 (35.37) | 28,088 (36.51) | 29,335 (36.46) |
| 21-30 | 894 (26.73) | 15,131 (19.62) | 953 (27.03) | 15,072 (19.59) | 16,025 (19.92) |
| 31-40 | 555 (16.6) | 8,285 (10.74) | 558 (15.83) | 8,282 (10.76) | 8,840 (10.99) |
| >40 | 361 (10.8) | 5,274 (6.84) | 364 (10.32) | 5,271 (6.85) | 5,635 (7.0) |
| *Missing* | 12 (0.36) | 184 (0.24) | 8 (0.23) | 188 (0.24) | 196 (0.24) |
| Pack-years of smoking (n, %) | | | | | |
| <10 | 88 (2.68) | 12,874 (17.04) | 86 (2.48) | 12,876 (17.08) | 12,962 (16.44) |
| 10-19 | 196 (5.97) | 14,648 (19.39) | 219 (6.33) | 14,625 (19.4) | 14,844 (18.83) |
| 20-29 | 320 (9.74) | 11,382 (15.06) | 356 (10.29) | 11,346 (15.05) | 11,702 (14.84) |
| 30-39 | 328 (9.98) | 9,850 (13.04) | 341 (9.85) | 9,837 (13.05) | 10,178 (12.91) |
| ≥40 | 2,353 (71.63) | 26,805 (35.48) | 2,459 (71.05) | 26,699 (35.42) | 29,158 (36.98) |
| *Missing* | 71 (2.12) | 1744 (2.26) | 73 (2.07) | 1742 (2.26) | 1815 (2.25) |
| Personal history of cancer (n, %) | 216 (6.44) | 3,480 (4.5) | 220 (6.23) | 3,476 (4.51) | 3,696 (4.58) |
| *Missing* | 0 (0.0) | 17 (0.02) | 0 (0.0) | 17 (0.02) | 17 (0.02) |
| COPD / Emphysema / Bronchitis (n, %) | 607 (18.09) | 6,615 (8.56) | 584 (16.53) | 6,638 (8.61) | 7,222 (8.95) |
| *Missing* | 0 (0.0) | 0 (0.0) | 0 (0.0) | 0 (0.0) | 0 (0.0) |
| Family history of lung cancer (n, %) | 562 (17.73) | 8,479 (11.39) | 573 (17.2) | 8,468 (11.4) | 9,041 (11.65) |
| *Missing* | 187 (5.57) | 2891 (3.74) | 202 (5.72) | 2876 (3.73) | 3078 (3.82) |

**eTable 9:** Outcomes by dataset

| Years | UK Biobank | | NLST control arm | | PLCO ever-smokers (intervention arm) | | PLCO ever-smokers (all) | |
|---|---|---|---|---|---|---|---|---|
| | Lung cancers (n) | Deaths from lung cancer (n) | Lung cancers (n) | Deaths from lung cancer (n) | Lung cancers (n) | Deaths from lung cancer (n) | Lung cancers (n) | Deaths from lung cancer (n) |
| 1 | 202 | 55 | 185 | 36 | 169 | 32 | 261 | 55 |
| 2 | 439 | 194 | 314 | 105 | 281 | 105 | 489 | 195 |
| 3 | 712 | 356 | 442 | 186 | 412 | 188 | 747 | 358 |
| 4 | 1010 | 548 | 572 | 276 | 543 | 271 | 993 | 534 |
| 5 | 1335 | 737 | 719 | 365 | 643 | 351 | 1231 | 727 |
| 6 | 1653 | 956 | 885 | 481 | 754 | 445 | 1464 | 914 |
| 7 | 2060 | 1189 | 957 | 545 | 915 | 538 | 1744 | 1106 |
| 8 | 2456 | 1463 | 959 | - | 1046 | 655 | 2019 | 1329 |
| 9 | 2790 | 1722 | - | - | 1184 | 777 | 2303 | 1566 |
| 10 | 3112 | 1930 | - | - | 1330 | 881 | 2573 | 1770 |

UK Biobank and the US National Lung Screening Trial were model development datasets. External validation occurred amongst ever-smokers in the PLCO intervention arm and amongst all ever-smokers in the PLCO.

**eTable 10:** Discrimination of UCL-D, Cox models, and the constrained LCDRAT, LCRAT, and PLCOm2012 models

| | Predictors | AUC – UK Biobank | AUC – PLCO radiography arm |
|---|---|---|---|
| UCL-D | 3 | 0.826 (0.815-0.838) | 0.803 (0.783-0.824) |
| Cox model (no interactions) | 3 | 0.817 (0.809-0.825) | 0.782 (0.772-0.793) |
| Cox model with interactions | 3 | 0.819 (0.811-0.827) | 0.785 (0.775-0.795) |
| LCDRAT-constrained | 6 | 0.821 (0.807-0.833) | 0.801 (0.782-0.820) |
| LCRAT-constrained | 6 | 0.806 (0.796-0.819) | 0.788 (0.773-0.803) |
| PLCOm2012-constrained | 5 | 0.786 (0.772-0.799) | 0.778 (0.766-0.797) |

UCL-D, the two Cox models, and LCDRAT-constrained predict 5-year risk of lung cancer death; LCRAT-constrained and PLCOm2012 constrained 5-year risk of lung cancer occurrence. Cox models were modelled with restricted cubic splines, with and without mutual interactions between age, smoking duration, and pack-years. The LCDRAT and LCRAT-constrained models use age, sex, quit-years, smoking duration, cigarettes per day, and pack-years. PLCOm2012-constrained uses age, smoking status, smoking duration, cigarettes per day, and quit-years. Both the LCRAT/LCDRAT and PLCOm2012 models were developed in the control arm of the PLCO trial. The relatively shallow drop-off in discriminatory performance between the various constrained models and their full versions show the relative importance of few smoking parameters and validates our findings that few smoking variables drive all lung cancer models in ever-smokers. The improvement seen by UCL-D over Cox models using the same data and variables reflects the statistical advantages of ensemble machine learning approaches.

**eTable 11:** Brier scores in UK Biobank and PLCO radiography arm

| | Risk of death from lung cancer | | Risk of developing lung cancer | | | |
|---|---|---|---|---|---|---|
| | UCL-D | LCDRAT | UCL-I | LCRAT | PLCOm2012* | LLPv2 |
| | | | **UK Biobank** | | | |
| Overall | 0.0034 (0.0031-0.0036) | 0.0034 (0.0031-0.0036) | 0.006 (0.0058-0.0064) | 0.006 (0.0057-0.0063) | 0.006 (0.0057-0.0063) | 0.0062 (0.0059-0.0065) |
| Age category | | | | | | |
| 40-49 | 0.0005 (0.0002-0.0007) | 0.0005 (0.0002-0.0007) | 0.0009 (0.0006-0.0012) | 0.0009 (0.0006-0.0013) | 0.0009 (0.0006-0.0013) | 0.0009 (0.0006-0.0013) |
| 50-59 | 0.0025 (0.0022-0.0029) | 0.0025 (0.0021-0.0028) | 0.0043 (0.0038-0.0047) | 0.0042 (0.0038-0.0048) | 0.0042 (0.0038-0.0048) | 0.0042 (0.0038-0.0049) |
| 60-72 | 0.0052 (0.0047-0.0056) | 0.0051 (0.0048-0.0056) | 0.0094 (0.0088-0.0100) | 0.0093 (0.0090-0.0099) | 0.0094 (0.0090-0.0099) | 0.0097 (0.0093-0.0102) |
| Sex | | | | | | |
| Female | 0.0029 (0.0026-0.0031) | 0.0028 (0.0025-0.0032) | 0.0056 (0.0051-0.0060) | 0.0055 (0.0050-0.0059) | 0.0055 (0.0050-0.0059) | 0.0056 (0.0050-0.0059) |
| Male | 0.0039 (0.0035-0.0042) | 0.0038 (0.0035-0.0042) | 0.0065 (0.0061-0.0070) | 0.0065 (0.0061-0.0069) | 0.0065 (0.0061-0.0069) | 0.0068 (0.0063-0.0072) |
| Smoking status | | | | | | |
| Former | 0.0023 (0.0020-0.0025) | 0.0022 (0.0020-0.0025) | 0.0043 (0.0041-0.0046) | 0.0043 (0.0040-0.0046) | 0.0043 (0.0040-0.0046) | 0.0044 (0.0042-0.0047) |
| Current | 0.0069 (0.0061-0.0076) | 0.0069 (0.0062-0.0075) | 0.0115 (0.0107-0.0124) | 0.0115 (0.0107-0.0123) | 0.0115 (0.0107-0.0123) | 0.0117 (0.0109-0.0125) |
| Ethnicity | | | | | | |
| Other | 0.0017 (0.0006-0.0023) | 0.0015 (0.0008-0.0023) | 0.0031 (0.0019-0.0042) | 0.0029 (0.0020-0.0041) | 0.0029 (0.0020-0.0040) | 0.0030 (0.0022-0.0042) |
| White | 0.0034 (0.0032-0.0037) | 0.0034 (0.0032-0.0037) | 0.0062 (0.0059-0.0065) | 0.0061 (0.0058-0.0064) | 0.0062 (0.0059-0.0065) | 0.0063 (0.0060-0.0066) |
| Household income | | | | | | |
| <18,000 | 0.0059 (0.0052-0.0064) | 0.0059 (0.0052-0.0065) | 0.0104 (0.0096-0.0112) | 0.0103 (0.0095-0.0112) | 0.0103 (0.0095-0.0113) | 0.0105 (0.0097-0.0115) |
| 18,000 to 30,999 | 0.0038 (0.0033-0.0042) | 0.0037 (0.0033-0.0042) | 0.0068 (0.0062-0.0074) | 0.0068 (0.0061-0.0075) | 0.0068 (0.0062-0.0075) | 0.0070 (0.0064-0.0077) |
| 31,000 to 51,999 | 0.0019 (0.0015-0.0021) | 0.0018 (0.0015-0.0022) | 0.0034 (0.0029-0.0039) | 0.0033 (0.0029-0.0039) | 0.0033 (0.0029-0.0039) | 0.0035 (0.0030-0.0040) |
| 52,000 to 100,000 | 0.0013 (0.0009-0.0016) | 0.0013 (0.0010-0.0017) | 0.0024 (0.0019-0.0029) | 0.0024 (0.0019-0.0029) | 0.0024 (0.0019-0.0029) | 0.0024 (0.0020-0.0030) |
| >100,000 | 0.0012 (0.0003-0.0017) | 0.0011 (0.0006-0.0018) | 0.0021 (0.0011-0.0029) | 0.0020 (0.0013-0.0031) | 0.0020 (0.0013-0.0031) | 0.0021 (0.0013-0.0032) |
| | | | **PLCO radiography arm** | | | |
| Overall | 0.0084 (0.0075-0.0093) | 0.0084 (0.0075-0.0093) | 0.0153 (0.0142-0.0164) | 0.0152 (0.0143-0.0164) | 0.0153 (0.0143-0.0164) | 0.0153 (0.0143-0.0165) |

| | | | | | | |
|---|---|---|---|---|---|---|
| Age category | | | | | | |
| 55-59 | 0.0046 (0.0037-0.0057) | 0.0046 (0.0036-0.0057) | 0.0090 (0.0077-0.0105) | 0.0090 (0.0077-0.0105) | 0.0090 (0.0077-0.0105) | 0.0091 (0.0078-0.0106) |
| 60-64 | 0.0068 (0.0059-0.0081) | 0.0069 (0.0059-0.0082) | 0.0127 (0.0110-0.0147) | 0.0128 (0.0111-0.0147) | 0.0128 (0.0111-0.0147) | 0.0128 (0.0110-0.0148) |
| 65-69 | 0.0122 (0.0100-0.0146) | 0.0122 (0.0100-0.0145) | 0.0224 (0.0198-0.0257) | 0.0223 (0.0197-0.0254) | 0.0222 (0.0197-0.0253) | 0.0225 (0.0199-0.0259) |
| 70-74 | 0.0160 (0.0129-0.0193) | 0.0161 (0.0129-0.0193) | 0.0261 (0.0219-0.0298) | 0.0263 (0.0221-0.0299) | 0.0265 (0.0226-0.0301) | 0.0263 (0.0220-0.0300) |
| Sex | | | | | | |
| Female | 0.0071 (0.0058-0.0083) | 0.0072 (0.0059-0.0084) | 0.0138 (0.0125-0.0155) | 0.0138 (0.0125-0.0155) | 0.0139 (0.0125-0.0155) | 0.0139 (0.0125-0.0156) |
| Male | 0.0093 (0.0082-0.0107) | 0.0093 (0.0082-0.0107) | 0.0163 (0.0150-0.0180) | 0.0162 (0.0149-0.0180) | 0.0163 (0.0150-0.0180) | 0.0163 (0.0151-0.0182) |
| Smoking status | | | | | | |
| Former | 0.0058 (0.0050-0.0066) | 0.0059 (0.0050-0.0066) | 0.0108 (0.0098-0.0118) | 0.0108 (0.0098-0.0119) | 0.0109 (0.0099-0.0119) | 0.0109 (0.0099-0.0119) |
| Current | 0.0187 (0.0160-0.0213) | 0.0187 (0.0160-0.0214) | 0.0329 (0.0300-0.0368) | 0.0329 (0.0299-0.0366) | 0.0330 (0.0300-0.0368) | 0.0331 (0.0301-0.0371) |
| Qualifications | | | | | | |
| Degree | 0.0060 (0.0047-0.0072) | 0.0060 (0.0047-0.0071) | 0.0109 (0.0092-0.0126) | 0.0109 (0.0091-0.0126) | 0.0109 (0.0091-0.0125) | 0.0110 (0.0092-0.0127) |
| Some college | 0.0076 (0.0061-0.0095) | 0.0076 (0.0061-0.0095) | 0.0144 (0.0122-0.0165) | 0.0144 (0.0122-0.0165) | 0.0144 (0.0122-0.0165) | 0.0145 (0.0123-0.0166) |
| Post-secondary | 0.0091 (0.0077-0.0105) | 0.0091 (0.0077-0.0105) | 0.0166 (0.0148-0.0187) | 0.0165 (0.0147-0.0187) | 0.0166 (0.0148-0.0187) | 0.0167 (0.0148-0.0189) |
| Secondary school | 0.0157 (0.0119-0.0200) | 0.0158 (0.0121-0.0201) | 0.0273 (0.0222-0.0324) | 0.0275 (0.0225-0.0325) | 0.0276 (0.0227-0.0324) | 0.0275 (0.0222-0.0326) |
| None of above | 0.0210 (0.0102-0.0391) | 0.0210 (0.0103-0.0387) | 0.0315 (0.0146-0.0492) | 0.0320 (0.0153-0.0496) | 0.0322 (0.0158-0.0494) | 0.0314 (0.0144-0.0490) |
| Ethnicity | | | | | | |
| Asian | 0.0069 (0.0031-0.0109) | 0.0070 (0.0030-0.0110) | 0.0111 (0.0056-0.0172) | 0.0111 (0.0056-0.0174) | 0.0112 (0.0057-0.0174) | 0.0112 (0.0057-0.0173) |
| Black | 0.0159 (0.0109-0.0209) | 0.0158 (0.0109-0.0205) | 0.0258 (0.0196-0.0316) | 0.0256 (0.0197-0.0313) | 0.0255 (0.0197-0.0309) | 0.0257 (0.0196-0.0316) |
| Other | 0.0106 (0.0058-0.0166) | 0.0105 (0.0057-0.0165) | 0.0146 (0.0091-0.0206) | 0.0146 (0.0090-0.0205) | 0.0150 (0.0095-0.0209) | 0.0145 (0.0089-0.0202) |
| White | 0.0079 (0.0070-0.0088) | 0.0079 (0.0070-0.0089) | 0.0148 (0.0136-0.0159) | 0.0147 (0.0135-0.0158) | 0.0148 (0.0136-0.0158) | 0.0149 (0.0137-0.0160) |

* The PLCOm2012 score predicts 6-year risk of developing lung cancer, whilst all other scores predict risk at 5-years. The Brier score varies by prevalence, so results presented here are for PLCOm2012 against 5-year outcomes to allow for direct comparison. Lower Brier scores indicate better model performance.

**eTable 12:** Model sensitivity and sensitivity at specified risk thresholds in the PLCO dataset

| | Risk threshold (%) | Sensitivity | Specificity |
|---|---|---|---|
| *Predicting 5-year risk of death from lung cancer* | | | |
| UCL-D | 0.68 | 0.855 (0.828-0.882) | 0.574 (0.570-0.577) |
| USPSTF-2021 | - | 0.775 (0.746-0.809) | 0.574 (0.570-0.578) |
| *Predicting 5-year risk of developing lung cancer* | | | |
| UCL-I | 1.17 | 0.839 (0.820-0.861) | 0.577 (0.574-0.580) |
| USPSTF-2021 | - | 0.777 (0.758-0.802) | 0.576 (0.572-0.579) |

Risk thresholds set using a fixed population approach at a level that would screen an equivalent number as the USPSTF-2021 in the PLCO external validation dataset. The entire PLCO dataset was used for these analyses.

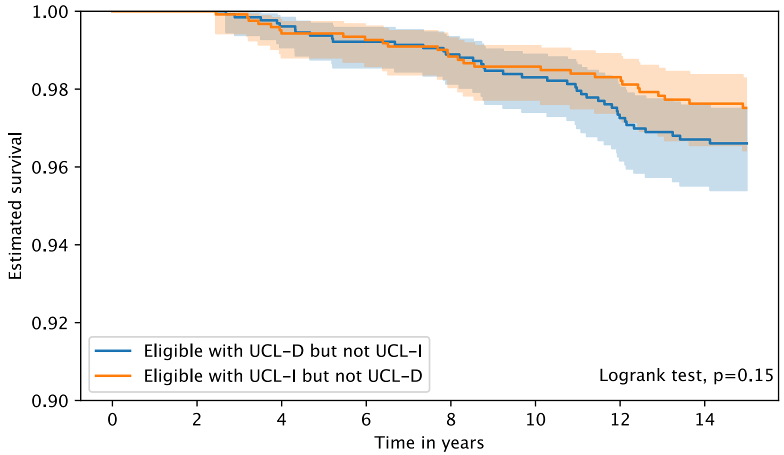

| | 0 | 2 | 4 | 6 | 8 | 10 | 12 | 14 |
|---|---|---|---|---|---|---|---|---|
| **Eligible with UCL-D but not UCL-I** | | | | | | | | |
| At risk | 1313 | 1299 | 1274 | 1239 | 1198 | 1150 | 1095 | 973 |
| Censored | 1 | 15 | 35 | 65 | 102 | 143 | 186 | 302 |
| Events | 0 | 0 | 5 | 10 | 14 | 21 | 33 | 39 |
| **Eligible with UCL-I but not UCL-D** | | | | | | | | |
| At risk | 1253 | 1245 | 1213 | 1186 | 1151 | 1110 | 1041 | 941 |
| Censored | 0 | 8 | 33 | 58 | 88 | 126 | 192 | 285 |
| Events | 0 | 0 | 7 | 9 | 14 | 17 | 20 | 27 |

(a)

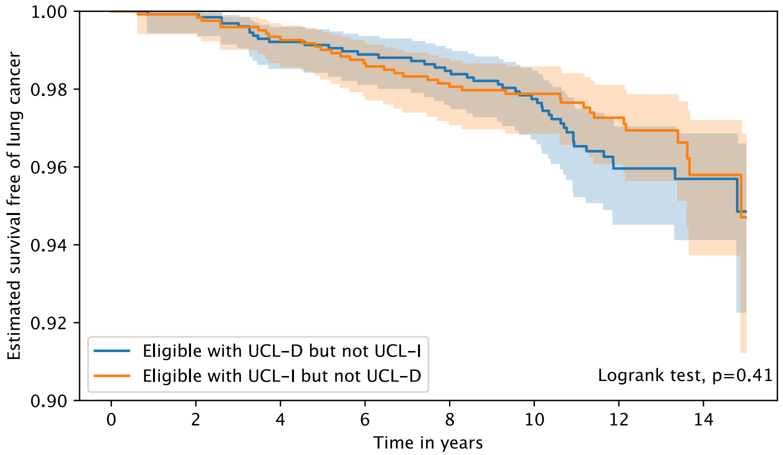

| | 0 | 2 | 4 | 6 | 8 | 10 | 12 | 14 |
|---|---|---|---|---|---|---|---|---|
| **Eligible with UCL-D but not UCL-I** | | | | | | | | |
| At risk | 1306 | 1284 | 1245 | 1204 | 1156 | 993 | 618 | 219 |
| Censored | 8 | 29 | 59 | 96 | 139 | 294 | 654 | 1052 |
| Events | 0 | 1 | 10 | 14 | 19 | 27 | 42 | 43 |
| **Eligible with UCL-I but not UCL-D** | | | | | | | | |
| At risk | 1246 | 1230 | 1191 | 1156 | 1113 | 971 | 622 | 182 |
| Censored | 7 | 22 | 53 | 81 | 117 | 257 | 601 | 1036 |
| Events | 0 | 1 | 9 | 16 | 23 | 25 | 30 | 35 |

(b)

**eFigure 9:** Outcomes by eligibility for either UCL-D or UCL-I, but not both UCL models

Kaplan-Meier plots showing lung cancer deaths (a) and lung cancers (b) amongst the 1,314 participants in the PLCO trial who would be eligible for screening with UCL-D (assuming a cut-off of 0.68%) but not UCL-I (assuming a cut-off of 1.17%) [blue lines] and amongst the 1,253 individuals who would be eligible for screening with UCL-I but not UCL-D [orange lines]. Overall, 34,756 individuals in the PLCO trial would have been eligible for screening with UCL-D at a cut-off of 0.68% and 34,695 with a cut-off of 1.17%. There is a trend shown in (a) towards those who would have been eligible for UCL-D but not UCL-I having a lower survival beyond 10 years (i.e., more deaths from lung cancer), but the difference did not reach statistical significance.

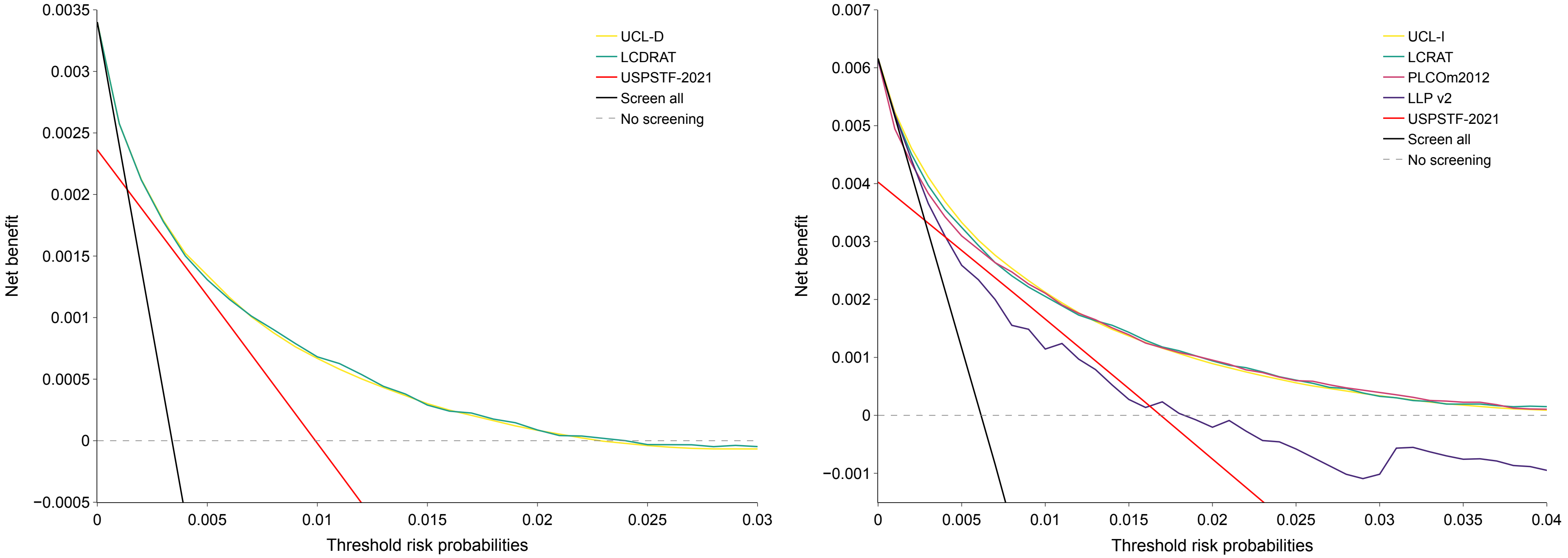

(a) (b)

**eFigure 10:** Net benefit of models in the UK Biobank

Net benefit across a range of thresholds of models predicting 5-year risk of death from lung cancer (A) and developing lung cancer (B) compared against USPSTF-2021 screening eligibility criteria in the UK Biobank dataset. Risk model-based approaches had a higher net benefit than screening either all men or using the USPSTF-2021 criteria to determine screening eligibility. The PLCOm2012 was originally fitted to predict 6-year risk of lung cancer; to make comparison possible, here net benefit was calculated based on predicting 5-year risk of lung cancer. The performance of PLCOm2012 over a 5-year timeframe was equivalent to that of a 6-year timeframe in the UK Biobank with an AUC of 0.796 (0.782-0.811) and a Brier score of 0.006 (0.0057-0.0063). In this analysis, the difference in net benefit between two approaches to selecting for screening at a particular risk threshold would be the change in true positives at that threshold level who would be screened given a fixed number of false positives.[27]

### Full Models

We present analyses in the full (control and radiography arm together) of the PLCO dataset. Here we also include the ‘full’ models developed alongside UCL-D and UCL. Both models are machine learning ensembles that were developed on the combined UK Biobank-NLST dataset.

#### UCLFull-D

UCLFull-D is an eight-variable model that predicts five-year risk of death from lung cancer with the following predictors:

1. Age,
2. Smoking duration (years),
3. Pack-years,
4. Smoking intensity (number of cigarettes per day),
5. Quit-years,
6. Body-mass index,
7. Chronic obstructive pulmonary disease (COPD), and
8. Family history of lung cancer.

UCLFull – D = $(0.143 * AdaBoost) + (0.714 * Logistic\ Regression) + (0.143 * LightGBM)$

#### UCLFull-I

UCLFull-I is a six-variable model that predicts five-year risk of developing lung cancer using the following predictors:

1. Age,
2. Smoking duration (years),
3. Pack-years,
4. Body-mass index,
5. COPD, and
6. family history of lung cancer.

UCLFull-I=

$$(0.391 * CatBoost) + (0.391 * Logistic\ Regression) + (0.087 * AdaBoost) + (0.130 * LightGBM)$$

### eTable 13: Discriminative accuracy (AUC) in the whole PLCO cohort

| | Risk of death from lung cancer | | | Risk of developing lung cancer | | | | | |
|---|---|---|---|---|---|---|---|---|---|
| | UCLFull-D | UCL-D | LCDRAT | UCLFull-I | UCL-I | LCRAT | PLCOm2012 | LLP v2 | LLP v3 |
| PLCO overall | 0.80 (0.78-0.81) | 0.79 (0.78-0.81) | 0.80 (0.79-0.82) | 0.79 (0.78-0.8) | 0.78 (0.77-0.79) | 0.79 (0.78-0.8) | 0.79 (0.78-0.8) | 0.74 (0.73-0.76) | 0.74 (0.73-0.76) |
| Age category | | | | | | | | | |
| 55-59 | 0.82 (0.78-0.84) | 0.8 (0.77-0.83) | 0.82 (0.78-0.85) | 0.81 (0.78-0.83) | 0.79 (0.77-0.82) | 0.81 (0.79-0.84) | 0.8 (0.78-0.82) | 0.74 (0.71-0.76) | 0.74 (0.71-0.77) |
| 60-64 | 0.79 (0.76-0.81) | 0.79 (0.75-0.81) | 0.79 (0.76-0.81) | 0.78 (0.76-0.80) | 0.77 (0.75-0.80) | 0.78 (0.76-0.80) | 0.78 (0.77-0.80) | 0.74 (0.71-0.76) | 0.72 (0.70-0.75) |
| 65-69 | 0.77 (0.74-0.80) | 0.76 (0.73-0.79) | 0.78 (0.75-0.80) | 0.77 (0.74-0.79) | 0.76 (0.74-0.78) | 0.78 (0.75-0.80) | 0.78 (0.76-0.80) | 0.72 (0.70-0.75) | 0.73 (0.70-0.75) |
| 70-74 | 0.73 (0.70-0.77) | 0.73 (0.70-0.77) | 0.74 (0.71-0.78) | 0.73 (0.70-0.76) | 0.73 (0.70-0.76) | 0.73 (0.71-0.76) | 0.74 (0.71-0.77) | 0.69 (0.66-0.72) | 0.68 (0.65-0.71) |
| Sex | | | | | | | | | |
| Female | 0.81 (0.79-0.83) | 0.80 (0.78-0.82) | 0.81 (0.79-0.84) | 0.80 (0.78-0.81) | 0.78 (0.76-0.8) | 0.80 (0.78-0.81) | 0.80 (0.78-0.81) | 0.74 (0.72-0.76) | 0.74 (0.72-0.76) |
| Male | 0.79 (0.77-0.81) | 0.78 (0.76-0.8) | 0.80 (0.78-0.82) | 0.79 (0.77-0.80) | 0.78 (0.77-0.8) | 0.79 (0.78-0.81) | 0.79 (0.78-0.81) | 0.75 (0.73-0.76) | 0.75 (0.73-0.76) |
| Smoking status | | | | | | | | | |
| Former | 0.81 (0.78-0.83) | 0.80 (0.78-0.82) | 0.81 (0.79-0.83) | 0.79 (0.77-0.81) | 0.79 (0.77-0.80) | 0.79 (0.78-0.81) | 0.79 (0.78-0.81) | 0.75 (0.73-0.77) | 0.74 (0.73-0.76) |
| Current | 0.69 (0.66-0.72) | 0.67 (0.64-0.70) | 0.70 (0.67-0.73) | 0.69 (0.67-0.71) | 0.67 (0.65-0.69) | 0.70 (0.68-0.72) | 0.69 (0.67-0.71) | 0.64 (0.61-0.66) | 0.65 (0.62-0.67) |
| Qualifications | | | | | | | | | |
| Degree | 0.83 (0.80-0.86) | 0.83 (0.80-0.86) | 0.84 (0.81-0.87) | 0.82 (0.79-0.84) | 0.81 (0.78-0.83) | 0.82 (0.8-0.84) | 0.82 (0.8-0.84) | 0.77 (0.74-0.79) | 0.77 (0.74-0.79) |
| Some college | 0.79 (0.76-0.82) | 0.79 (0.75-0.82) | 0.78 (0.75-0.81) | 0.79 (0.77-0.82) | 0.79 (0.77-0.81) | 0.79 (0.76-0.81) | 0.80 (0.77-0.82) | 0.75 (0.73-0.78) | 0.74 (0.72-0.77) |
| Post-secondary | 0.79 (0.77-0.82) | 0.78 (0.75-0.80) | 0.79 (0.77-0.81) | 0.78 (0.76-0.80) | 0.77 (0.75-0.79) | 0.78 (0.76-0.8) | 0.77 (0.76-0.79) | 0.73 (0.71-0.75) | 0.73 (0.71-0.75) |
| Secondary school | 0.72 (0.67-0.78) | 0.71 (0.66-0.77) | 0.74 (0.68-0.78) | 0.72 (0.67-0.76) | 0.72 (0.67-0.76) | 0.74 (0.69-0.77) | 0.74 (0.70-0.77) | 0.68 (0.63-0.72) | 0.68 (0.63-0.72) |
| None of above | 0.68 (0.57-0.78) | 0.66 (0.53-0.76) | 0.70 (0.58-0.81) | 0.67 (0.56-0.75) | 0.66 (0.57-0.75) | 0.67 (0.55-0.76) | 0.68 (0.58-0.77) | 0.61 (0.51-0.73) | 0.62 (0.51-0.74) |
| Ethnicity | | | | | | | | | |
| Asian | 0.85 (0.79-0.91) | 0.86 (0.80-0.91) | 0.83 (0.76-0.90) | 0.74 (0.64-0.84) | 0.74 (0.62-0.84) | 0.74 (0.66-0.83) | 0.75 (0.67-0.82) | 0.71 (0.61-0.81) | 0.70 (0.59-0.81) |
| Black | 0.79 (0.73-0.85) | 0.78 (0.72-0.84) | 0.81 (0.75-0.86) | 0.78 (0.72-0.83) | 0.77 (0.72-0.82) | 0.79 (0.74-0.83) | 0.79 (0.75-0.83) | 0.75 (0.71-0.80) | 0.74 (0.70-0.79) |
| Other | 0.81 (0.72-0.90) | 0.78 (0.67-0.87) | 0.83 (0.76-0.91) | 0.80 (0.74-0.86) | 0.78 (0.71-0.85) | 0.81 (0.74-0.87) | 0.78 (0.70-0.84) | 0.73 (0.64-0.83) | 0.73 (0.64-0.83) |
| White | 0.80 (0.78-0.81) | 0.79 (0.78-0.80) | 0.80 (0.78-0.81) | 0.79 (0.78-0.80) | 0.78 (0.77-0.79) | 0.79 (0.78-0.81) | 0.80 (0.79-0.81) | 0.74 (0.73-0.76) | 0.74 (0.73-0.76) |

Note that the LCDRAT, LCRAT, and PLCOm2012 models were developed in the control arm of the PLCO cohort. The relative performance of the UCL models is therefore notable.

### eTable 14: Overall performance (Brier scores) in the whole PLCO cohort

| | Risk of death from lung cancer | | | Risk of developing lung cancer | | | | | |
|---|---|---|---|---|---|---|---|---|---|
| | UCLFull-D | UCL-D | LCDRAT | UCLFull-I | UCL-I | LCRAT | PLCOm2012* | LLP v2 | LLP v3 |
| PLCO overall | 0.0088 (0.0083-0.0094) | 0.0089 (0.0083-0.0094) | 0.0088 (0.0083-0.0094) | 0.0147 (0.014-0.0155) | 0.0147 (0.0141-0.0155) | 0.0147 (0.0141-0.0155) | 0.0147 (0.0141-0.0155) | 0.0148 (0.0142-0.0156) | 0.0149 (0.0143-0.0157) |
| Age category | | | | | | | | | |
| 55-59 | 0.0048 (0.0042-0.0056) | 0.0048 (0.0042-0.0056) | 0.0048 (0.0041-0.0055) | 0.0087 (0.0077-0.0099) | 0.0087 (0.0077-0.0099) | 0.0086 (0.0077-0.0098) | 0.0087 (0.0077-0.0098) | 0.0088 (0.0077-0.01) | 0.0088 (0.0078-0.01) |
| 60-64 | 0.0074 (0.0063-0.0082) | 0.0074 (0.0063-0.0082) | 0.0074 (0.0063-0.0082) | 0.0128 (0.0114-0.0139) | 0.0128 (0.0115-0.014) | 0.0128 (0.0115-0.014) | 0.0128 (0.0115-0.014) | 0.0128 (0.0115-0.014) | 0.0129 (0.0115-0.0141) |
| 65-69 | 0.0127 (0.0113-0.0143) | 0.0127 (0.0113-0.0144) | 0.0127 (0.0113-0.0143) | 0.0207 (0.019-0.0225) | 0.0208 (0.0191-0.0225) | 0.0207 (0.019-0.0224) | 0.0207 (0.019-0.0224) | 0.021 (0.0192-0.0228) | 0.0212 (0.0193-0.023) |
| 70-74 | 0.0166 (0.0146-0.0192) | 0.0167 (0.0146-0.0192) | 0.0166 (0.0146-0.0192) | 0.0254 (0.0225-0.0279) | 0.0255 (0.0225-0.028) | 0.0255 (0.0226-0.0281) | 0.0258 (0.0228-0.0283) | 0.0256 (0.0227-0.0281) | 0.0258 (0.0227-0.0284) |
| Sex | | | | | | | | | |
| Female | 0.0072 (0.0065-0.0081) | 0.0072 (0.0065-0.0081) | 0.0073 (0.0065-0.0081) | 0.013 (0.0119-0.0141) | 0.013 (0.012-0.0141) | 0.013 (0.0119-0.0141) | 0.0131 (0.012-0.0142) | 0.0131 (0.0121-0.0142) | 0.0131 (0.0121-0.0143) |
| Male | 0.0099 (0.009-0.0109) | 0.0099 (0.009-0.0109) | 0.0099 (0.009-0.0109) | 0.0159 (0.0146-0.017) | 0.0159 (0.0146-0.017) | 0.0159 (0.0146-0.017) | 0.0159 (0.0146-0.017) | 0.016 (0.0147-0.0171) | 0.0161 (0.0148-0.0173) |
| Smoking status | | | | | | | | | |
| Former | 0.0062 (0.0057-0.0068) | 0.0062 (0.0057-0.0068) | 0.0062 (0.0057-0.0068) | 0.0104 (0.0096-0.0111) | 0.0104 (0.0096-0.0111) | 0.0104 (0.0096-0.0111) | 0.0104 (0.0096-0.0111) | 0.0104 (0.0096-0.0111) | 0.0104 (0.0097-0.0111) |
| Current | 0.0193 (0.0172-0.021) | 0.0194 (0.0173-0.0211) | 0.0194 (0.0173-0.021) | 0.0322 (0.0298-0.0344) | 0.0324 (0.03-0.0346) | 0.0323 (0.0298-0.0344) | 0.0324 (0.03-0.0346) | 0.0326 (0.0301-0.0349) | 0.033 (0.0304-0.0354) |
| Qualifications | | | | | | | | | |
| Degree | 0.0059 (0.0051-0.0068) | 0.0059 (0.0051-0.0069) | 0.0059 (0.0051-0.0069) | 0.0104 (0.0094-0.0116) | 0.0104 (0.0094-0.0117) | 0.0104 (0.0094-0.0116) | 0.0104 (0.0094-0.0116) | 0.0105 (0.0095-0.0117) | 0.0105 (0.0095-0.0118) |
| Some college | 0.0083 (0.0072-0.0094) | 0.0083 (0.0072-0.0094) | 0.0083 (0.0072-0.0094) | 0.0143 (0.0126-0.0161) | 0.0144 (0.0126-0.0161) | 0.0144 (0.0127-0.0161) | 0.0144 (0.0127-0.0161) | 0.0145 (0.0127-0.0162) | 0.0146 (0.0128-0.0164) |
| Post-secondary | 0.0099 (0.0088-0.0112) | 0.01 (0.0089-0.0113) | 0.0099 (0.0088-0.0112) | 0.0162 (0.015-0.0177) | 0.0163 (0.015-0.0178) | 0.0162 (0.015-0.0176) | 0.0163 (0.015-0.0177) | 0.0164 (0.0151-0.0179) | 0.0165 (0.0152-0.018) |
| Secondary school | 0.0161 (0.0131-0.0189) | 0.016 (0.0131-0.019) | 0.0161 (0.0132-0.0189) | 0.0254 (0.0213-0.0287) | 0.0253 (0.0212-0.0286) | 0.0254 (0.0214-0.0286) | 0.0254 (0.0215-0.0286) | 0.0255 (0.0214-0.0288) | 0.0258 (0.0215-0.0292) |
| None of above | 0.0197 (0.0121-0.0301) | 0.0197 (0.0122-0.0302) | 0.0196 (0.0121-0.0299) | 0.0243 (0.0164-0.0366) | 0.0243 (0.0163-0.0367) | 0.0248 (0.017-0.0372) | 0.025 (0.0175-0.0374) | 0.0243 (0.0163-0.0367) | 0.0244 (0.0162-0.037) |

| | Risk of death from lung cancer | | | Risk of developing lung cancer | | | | | |
|---|---|---|---|---|---|---|---|---|---|
| | UCLFull-D | UCL-D | LCDRAT | UCLFull-I | UCL-I | LCRAT | PLCOm2012* | LLP v2 | LLP v3 |
| Ethnicity | | | | | | | | | |
| Asian | 0.0054 (0.0029-0.0079) | 0.0054 (0.0028-0.0079) | 0.0054 (0.0028-0.008) | 0.0089 (0.0057-0.0131) | 0.0089 (0.0057-0.0131) | 0.0088 (0.0056-0.0132) | 0.0089 (0.0056-0.0132) | 0.009 (0.0057-0.0132) | 0.0089 (0.0056-0.0132) |
| Black | 0.0143 (0.0108-0.0177) | 0.0144 (0.0109-0.0178) | 0.0141 (0.0107-0.0175) | 0.0222 (0.0179-0.0261) | 0.0223 (0.0179-0.0261) | 0.022 (0.0178-0.0256) | 0.022 (0.0179-0.0256) | 0.0224 (0.018-0.0263) | 0.0227 (0.0183-0.0267) |
| Other | 0.0077 (0.0043-0.0111) | 0.0077 (0.0042-0.0112) | 0.0077 (0.0042-0.0111) | 0.0114 (0.0076-0.0151) | 0.0114 (0.0077-0.0151) | 0.0114 (0.0076-0.0151) | 0.0117 (0.008-0.0155) | 0.0114 (0.0077-0.0151) | 0.0115 (0.0076-0.0153) |
| White | 0.0086(0.008-0.0092) | 0.0086 (0.008-0.0092) | 0.0086 (0.008-0.0092) | 0.0145 (0.0138-0.0153) | 0.0146 (0.0138-0.0154) | 0.0146 (0.0138-0.0154) | 0.0146 (0.0139-0.0154) | 0.0147 (0.0139-0.0155) | 0.0148 (0.014-0.0156) |

Note that the LCDRAT, LCRAT, and PLCOm2012 models were developed in the control arm of the PLCO cohort.
The PLCOm2012 was originally developed to predict 6-year risk of developing lung cancer. As Brier scores depend on prevalence, we present results for the PLCOm2012 against 5-year outcomes for the purposes of comparison

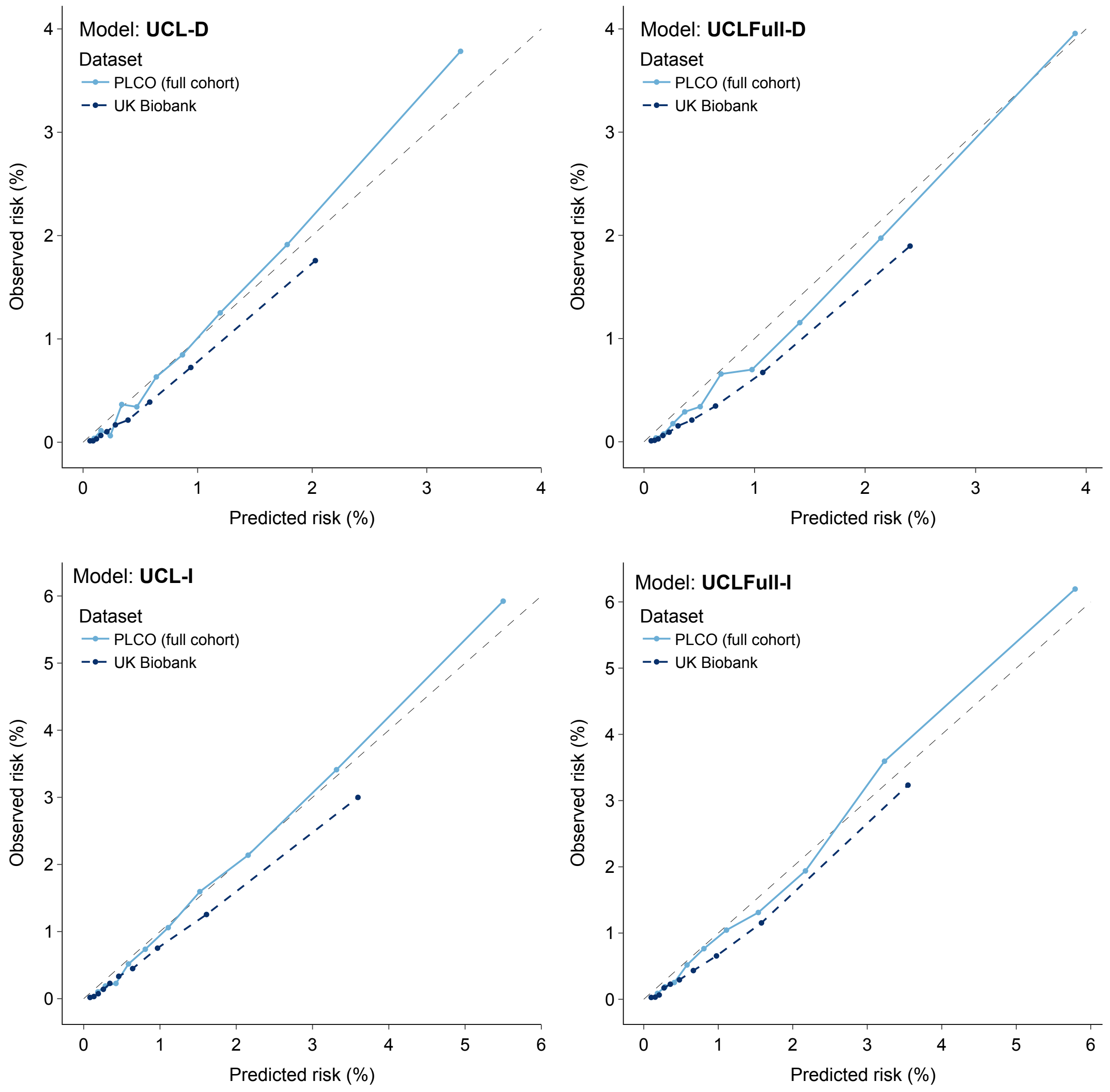

**eFigure 12:** Calibration curves for UCL models in the full PLCO cohort

Calibration curves showing observed against predicted risks in the PLCO full cohort (light blue) and UK Biobank (dark blue).

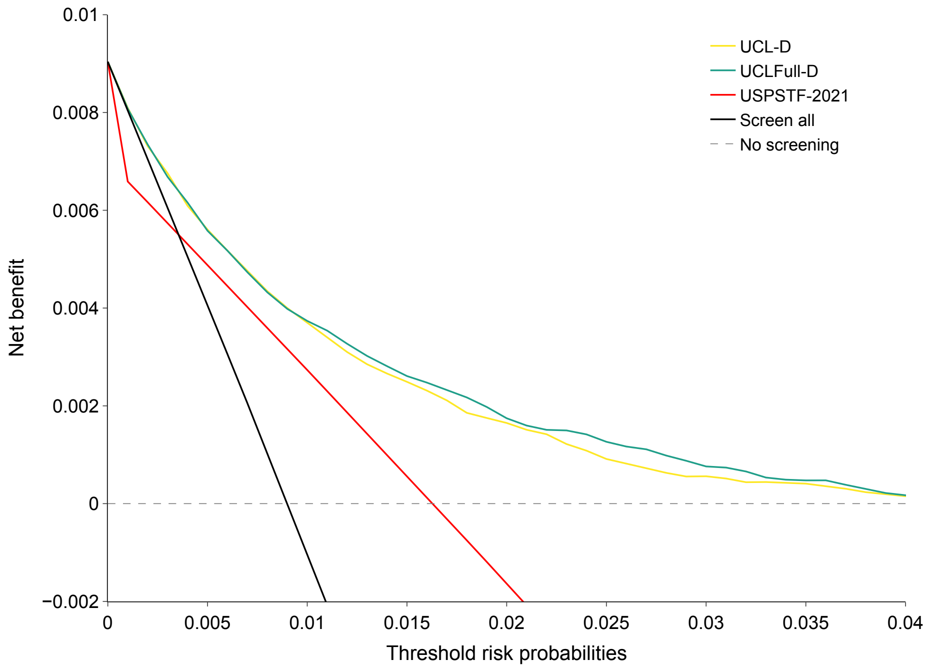

(a)

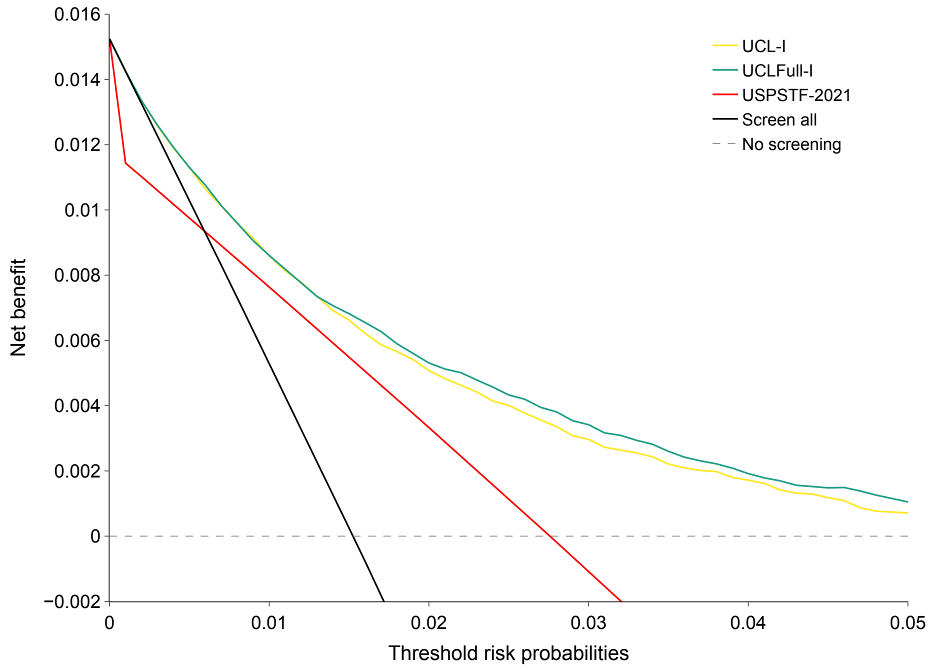

(b)

**eFigure 13:** Net benefit of UCL models in the full PLCO cohort

Net benefit across a range of thresholds of models predicting 5-year risk of death from lung cancer (a) and developing lung cancer (b) compared against USPSTF-2021 screening eligibility criteria in the PLCO cohort.

**eTable 15:** Sensitivity and sensitivity of UCLFull-D and UCLFull-I at specified risk thresholds in the PLCO dataset

| | Risk threshold (%) | Sensitivity | Specificity |
|---|---|---|---|
| *Predicting 5-year risk of death from lung cancer* | | | |
| UCLFull-D | 0.74 | 0.849 (0.821-0.874) | 0.574 (0.571-0.578) |
| USPSTF-2021 | - | 0.775 (0.746-0.809) | 0.574 (0.570-0.578) |
| *Predicting 5-year risk of developing lung cancer* | | | |
| UCLFull-I | 1.18 | 0.843 (0.822-0.863) | 0.577 (0.573-0.580) |
| USPSTF-2021 | - | 0.777 (0.758-0.802) | 0.576 (0.572-0.579) |

Risk thresholds set using a fixed population approach at a level that would screen an equivalent number as the USPSTF-2021 in the entire PLCO dataset.

10. Scikit-learn. An AdaBoost Classifier. https://scikit-learn.org/stable/modules/generated/sklearn.ensemble.AdaBoostClassifier.html#sklearn.ensemble.AdaBoostClassifier

11. Pedregosa F, Varoquaux G, Gramfort A, et al. Scikit-learn: Machine Learning in Python. *J Mach Learn Res*.

12. Prokhorenkova L, Gusev G, Vorobev A, Dorogush AV, Gulin A. CatBoost: unbiased boosting with categorical features. In: Bengio S, Wallach H, Larochelle H, Grauman K, Cesa-Bianchi N, Garnett R, eds. *Advances in Neural Information Processing Systems*. Vol 31. Curran Associates, Inc.; 2018. https://proceedings.neurips.cc/paper/2018/file/14491b756b3a51daac41c24863285549-Paper.pdf

13. Yandex. CatBoost. https://catboost.ai/en/docs/

14. Ke G, Meng Q, Finley T, et al. LightGBM: A highly efficient gradient boosting decision tree. *Adv Neural Inf Process Syst*. 2017;30. Accessed January 10, 2023. https://proceedings.neurips.cc/paper/2017/hash/6449f44a102fde848669bdd9eb6b76fa-

Abstract.html

15. Microsoft. LightGBM. https://lightgbm.readthedocs.io/en/v3.3.4/index.html

16. Chen T, Guestrin C. XGBoost: A Scalable Tree Boosting System. *arXiv [csLG]*. Published online 9 March 2016. http://arxiv.org/abs/1603.02754

17. XGBoost. https://xgboost.readthedocs.io/en/stable/index.html

18. Akiba T, Sano S, Yanase T, Ohta T, Koyama M. Optuna: A Next-generation Hyperparameter Optimization Framework. In: *Proceedings of the 25th ACM SIGKDD International Conference on Knowledge Discovery & Data Mining*. KDD ’19. Association for Computing Machinery; 2019:2623-2631. doi:10.1145/3292500.3330701

19. Zhao Y, Wang X, Cheng C, Ding X. Combining Machine Learning Models and Scores using combo library. In: *Thirty-Fourth AAAI Conference on Artificial Intelligence*. ; 2020.

20. Therneau TM. *A Package for Survival Analysis in R*. https://CRAN.R-project.org/package=survival

21. Harrel FE. *Regression Modelling Strategies*. https://cran.r-project.org/web/packages/rms/index.html
